# Investigating the impact of sex and reproductive aging on latent signatures of modifiable dementia risk factors

**DOI:** 10.1101/2025.02.21.25322638

**Authors:** Alice Mukora, Manuela Costantino, Olivier Parent, Gabriel. A. Devenyi, M. Mallar Chakravarty

## Abstract

Treating modifiable risk factors for dementia may serve to prevent maladaptive brain aging. These include, and are not limited to, obesity, hypertension, and other metabolic disorders. Nearly two-thirds of affected individuals are female, and emerging research has pointed to potential sex-specific factors, like reproductive aging, as potent modifiers of dementia risk. Here, we leverage neuroimaging to characterize sex differences in the neural signatures of modifiable dementia risk factors, examining their relationship with brain anatomy, and integrate female-specific factors related to menopause history in a sex-specific analysis. Using cross-sectional data from the UK Biobank, we selected a cohort of 31,711 (age 44-82, 55.2% female/44.8% male) participants without diagnosed neurological conditions and collated behavioral data previously determined as modifiable risk factors. Using Partial Least Squares Analysis (PLS), we examined latent signatures that represent linear combinations that maximize covariance between patterns of brain (mean cortical thickness from 64 regions) and risk factor variables. To examine sex differences, we performed PLS analysis using the entire sample. We performed linear models to explore age-by-sex interactions with the PLS-derived brain scores and the risk factor pattern scores. To examine sex-specific relationships, we performed separate PLS analyses for the males and females and integrated menopause-related variables into the latter analysis. Our study found sex-dependent and menopause-dependent relationships between lifestyle risk factors and cortical thickness, highlighting stronger impacts of cardiometabolic factors on males and social and obesity-related factors on females in preserving brain health. The inclusion of menopause-related variables did not change the relationships to lifestyle risk factors, and strict age-matching dampened the strength of the findings. Our findings suggest that lifestyles and the female-specific endocrine environment influence sex differences in cortical anatomy during brain aging.

## 1. Introduction

As the global population continues to age, public health concerns related to disease and maladaptive aging are on the rise (Government of Canada, 2022; Nichols et al., 2022). One such concern is dementia, a family of progressive neurodegenerative disorders that impair cognitive function. Despite extensive research, current treatments for dementia have not been effective in halting or slowing the progression of disease, and no cure is currently available (Fessel, 2023). This has prompted investigation into modifiable risk factors that may serve as a promising arena to delay or prevent maladaptive neurodegeneration. A variety of risk factors, including obesity, hypertension, and smoking, have been found to account for 40% of global incidence (Livingston et al., 2020) and may serve as effective targets for risk reduction (Grodstein et al., 2023). While age is the primary risk factor for dementia, women are disproportionately affected by dementia, accounting for nearly two-thirds of affected individuals and having double the lifetime risk compared to men (Alzheimer’s Association, 2023). Historically, this higher incidence has been attributed to women’s longer life expectancy relative to men. However, emerging research suggests sex-dependent associations of risk factors with dementia incidence, cognitive decline, and neurodegeneration (Conde et al., 2021; Huo et al., 2022; Rocca et al., 2014). Profiles of dementia risk may be different between males and females due to sex differences in the prevalence of behaviors and environmental factors (Xiong et al., 2024), such as higher educational attainment and alcohol consumption (White, 2020; Wilsnack & Wilsnack, 2013) in older men. Sex also modulates the interactions with risk factors that have been associated with accelerated brain aging in women, such as waist-to-hip ratio (WHR) and body mass index (BMI) (Subramaniapillai et al., 2022, 2024); in contrast, lower educational attainment and former alcohol consumption have stronger associations with dementia incidence in men (Gong et al., 2023).

In addition to the above-mentioned sex-lifestyle interactions, women also possess unique risk factors, including those tied to reproductive history and reproductive aging (Azad et al., 2007; Than et al., 2022; Xiong et al., 2024). The menopause transition (MT), a neuroendocrine aging process specific to females, is understudied in this context. The MT has been linked to alterations of brain structure, particularly in areas involved in higher cognitive processes, though some studies have suggested those changes stabilize post-menopause (Mosconi et al., 2021; Ramli et al., 2023). Early timing of the menopause onset, either through spontaneous or surgical means, has been associated with increased risk for cognitive decline (Liao et al., 2023). Other endogenous or exogenous alterations to sex-hormone concentrations, such as pregnancy and the use of menopausal hormone therapy (MHT), can also negatively impact cognitive function and increase dementia risk in later life (Barth et al., 2024; Barth & de Lange, 2020; Crestol et al., 2024). In attempts to account for the inherent age differences between menopausal groups, studies from our group have utilized age-matched menopause groups and have found attenuated or undetectable differences in neuroanatomy and cognition between menopausal groups (Costantino et al., 2023; Wezel et al., 2024). These findings suggested that while menopause-specific changes may play a role in dementia risk, careful consideration should be taken in isolating the effect of the menopausal transition from the effect of aging.

In this paper, we use a large sample from the UK Biobank to investigate the associations between modifiable dementia risk factors and brain anatomy in mid to late life, both at a population level and with a sex-specific focus. Furthermore, in females, we examine how these associations vary with menopause status and type. We leveraged neuroimaging-derived cortical thickness measures to non-invasively quantify brain morphology and investigated associations with a wide array of dementia risk factors, aiming to identify distinct anatomical patterns linked to risk factor profiles across chronological aging and changes in the reproductive system.

## 2. Materials and Methods

### 2.1. Sample

Cross-sectional data was obtained from the UK Biobank, a large-scale open-access multimodal biomedical database (Application #45551). Full details of recruitment and other key UK Biobank procedures have been published and are available on www.ukbiobank.ac.uk. All participants provided written informed consent. An extensive magnetic resonance imaging (MRI) protocol was acquired for ∼40,000 participants between 2014 and 2020. Extensive data related to health and lifestyle was also acquired. We selected a cohort of 31,711 participants (age 44-82, 55.2% female/44.8% male) (**Table 1**) with available imaging data and passed the quality control metrics outlined below. 14,428 female participants had complete data related to the reproductive aging analyses.

**Table 1:** Basic demographic information for the participants in each analysis.

|  | N | Age (years)* |
| --- | --- | --- |
| **Sex Differences** |  |  |
| Females | 17,594 | 63.08 (7.03) |
| Males | 14,661 | 63.88 (7.58) |
| **Sex Specific** |  |  |
| Females | 14,428 | 63.28 (6.72) |
| Males | 14,208 | 63.82 (7.57) |
| **Menopause** |  |  |
| Premenopausal | 222 | 53.42 (2.39) |
| Natural Menopause | 445 | 53.78 (2.67) |
| Surgical Menopause | 223 | 54.11 (2.95) |
| Age-Matched Males | 890 | 53.00 (2.64) |

\*Mean (Standard Deviation)

### 2.2. Image Processing

UK Biobank used a standard 3T Siemens Skyra with a standard Siemens 32-channel RF receive head coil across three dedicated imaging centers in the United Kingdom (Miller et al., 2016). We performed manual quality control for motion on the T1-weighted (MPRAGE, 1mm^3^) scans before image processing to avoid biases in morphometric measurements in subsets of the population, a process developed by our group (https://github.com/CoBrALab/documentation/wiki/Motion-Quality-Control-(QC)-Manual) (Bedford et al., 2020). The passed scans were input into CIVET 2.1.1, a cortical thickness estimation pipeline developed for human magnetic resonance (MR) images (Ad-Dab’bagh et al., 2006). Each scan was registered from native to stereotaxic space, using the MNI ICBM152 model as the registration target. Next, CIVET performed tissue classification into white matter (WM), gray matter (GM) and cerebrospinal fluid (CSF) based on the T1w image. WM and pial surfaces composed of 40,962 vertices were extracted for each hemisphere. Cortical thickness was calculated in mm as the distance between the pial and WM surfaces in the native space of the original image. Regional average cortical thickness values were then derived using the Desikan-Killiany-Tourville (DKT) atlas (Desikan et al., 2006; Klein & Tourville, 2012) applied to imaging data smoothed with a 30 mm full width at half maximum (*fwhm*) kernel. Finally, all cortical thickness outputs underwent a manual quality control step to ensure accurate tissue classification (https://github.com/CoBrALab/documentation/wiki/CIVET-Quality-Control-Guidelines).

As the project aims to explore healthy aging, participants with confounding neurological, psychiatric, and substance-related diagnoses that are known to impact neuroanatomy (n = 852) were excluded from analyses. Exposures were derived from self-report of illness or disability (UK Biobank Data Field 20002), and specific codes used for exclusion are detailed in **Supplementary Table 1.**

### 2.3. Lifestyle Variables

At each visit to the assessment center, participants provided information through questionnaires about alcohol, total household income, smoking, hearing difficulty, physical activity, sleep, social support, and social isolation. We reviewed the literature to identify available variables that aligned with the modifiable dementia risk factors outlined in the 2020 Lancet Commission report, covering cardiometabolic risk (hypertension, diabetes, smoking, obesity, high alcohol consumption, and physical inactivity), behavioral factors (depression, low educational attainment, low social contact), and other conditions (hearing impairment) (Livingston et al., 2020). Additionally, we conducted a manual keyword search to identify further relevant variables of interest that have been related to dementia risk, including sleep disturbances (Benito-León et al., 2009; Lutsey et al., 2018; Sindi et al., 2018) and markers of socioeconomic status (Hunt et al., 2020; Kivimäki et al., 2020; Livingston et al., 2020). All continuous variables were z-scored to obtain standardized beta coefficients. Results were corrected with the False Discovery Rate (FDR) (Benjamini & Hochberg, 1995) and are presented with a 5% FDR correction. We then assessed their relevance and alignment with the domains and life course stages associated with each behavior implicated in dementia risk. For instance, excessive alcohol consumption is considered a midlife risk factor (Anttila et al., 2004; Strandberg et al., 2018), so we chose to assess current alcohol consumption status and frequency, while discarding information about previous alcohol consumption and abstention. Preference was given to answers provided during the imaging visit. Variables with insufficient variance (i.e.: heavily skewed to one specific response category), as determined by qualitative assessment, or those missing data for more than 10% of participants were excluded. Any participant missing data for more than 10 demographic variables (n = 218) was excluded from analyses. Where appropriate, missing data was backfilled from visits preceding the imaging visit (n = 12,756).

Any remaining missing data was imputed using the missForest package in R 4.2.2. The categories used in our analyses were as reported in the UK Biobank questionnaire, unless otherwise noted. **Supplementary Table 2** contains the original names and codes of the fields from which all variables were derived.

### 2.4. Female-specific variables

Based on self-report data, female participants were divided into premenopausal women (PRE), women who underwent a bilateral oophorectomy (with or without a hysterectomy) prior to menopause (SURG) and women who underwent menopause without surgical interference (POST). Menopause status was assessed by self-report at each imaging visit, indicated as an answer of “Yes” or “No” to “Have you had your menopause (periods stopped)?” Any participant who reported an age at menopause at the imaging timepoint or any previous visit to an assessment center or was over 70 years old was coded as postmenopausal. Surgical menopausal status was determined by self-report of bilateral oophorectomy (with or without a hysterectomy) prior to menopause (SURG) and women who underwent menopause without surgical interference (POST). Menopause status was assessed by self-report at the imaging visit, indicated as an answer of “Yes” or “No” to “Have you had your menopause (periods stopped)?” Any participant who reported an age at menopause at the imaging timepoint or any previous visit to an assessment center or was over 70 years old was coded as postmenopausal. Surgical menopausal status was determined by self-report of bilateral oophorectomy and reported age at bilateral oophorectomy being at or prior to reported age at menopause. Any participant with uncertain menopause status or hysterectomy without a bilateral oophorectomy was excluded from any female-specific analysis. Binary (yes/no) variables were generated for analysis, encoding menopause status (premenopausal or postmenopausal), surgical menopause, or natural menopause.

### 2.5. Statistical Analysis

#### 2.5.1. Group Differences

We tested the interaction of female sex and age in cortical thickness measures in the whole sample (Eq 1) using vertex-wise linear models in R/4.2.2:

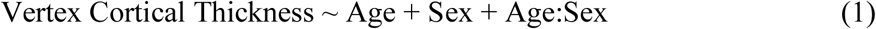

All continuous variables were z-scored to obtain standardized beta coefficients. Results were corrected with the False Discovery Rate (FDR) (Benjamini and Hochberg, 1995) and are presented with a 5% FDR correction.

To investigate group-level differences in neuroanatomy in the female sample, we used a vertex-wise analysis of variance model to test the interaction of age and menopause status (Eq 2).

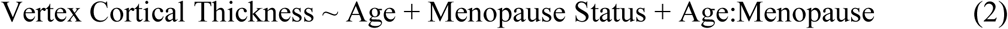

Results were FDR-corrected and presented with a 20% FDR threshold owing to the limited findings in this analysis.

#### 2.5.2. Partial Least Squares Analysis (PLS)

In order to assess the multivariate relationships between cortical thickness measures and risk factor behaviors, we performed partial least square (PLS) analysis, a technique that extracts patterns of covariance between two sets of variables (McIntosh & Lobaugh, 2004; McIntosh & Mišić, 2013) (**Figure 1**). The variables used were the brain matrix containing regional cortical thickness at each DKT atlas ROI, and the risk factor data matrix across all analysis-specific grouping of participants. These two matrices were standardized (z-scored) across each column and the covariance matrix was computed. This matrix was subjected to singular value decomposition, which yielded a set of latent variables (LV) - linear combinations of the variables in the brain and behavioral variables that maximally covary with one another (Zeighami et al., 2019). For each LV, each participant has a calculated brain and behavior score, defined as the singular value obtained from the decomposition of the respective matrix and is representative of the extent to which the participant expresses the cortical thickness and risk factor patterns of the LV. All analyses were performed using Pyls, a Python implementation of PLS (https://github.com/rmarkello/pyls).

**Figure 1:**
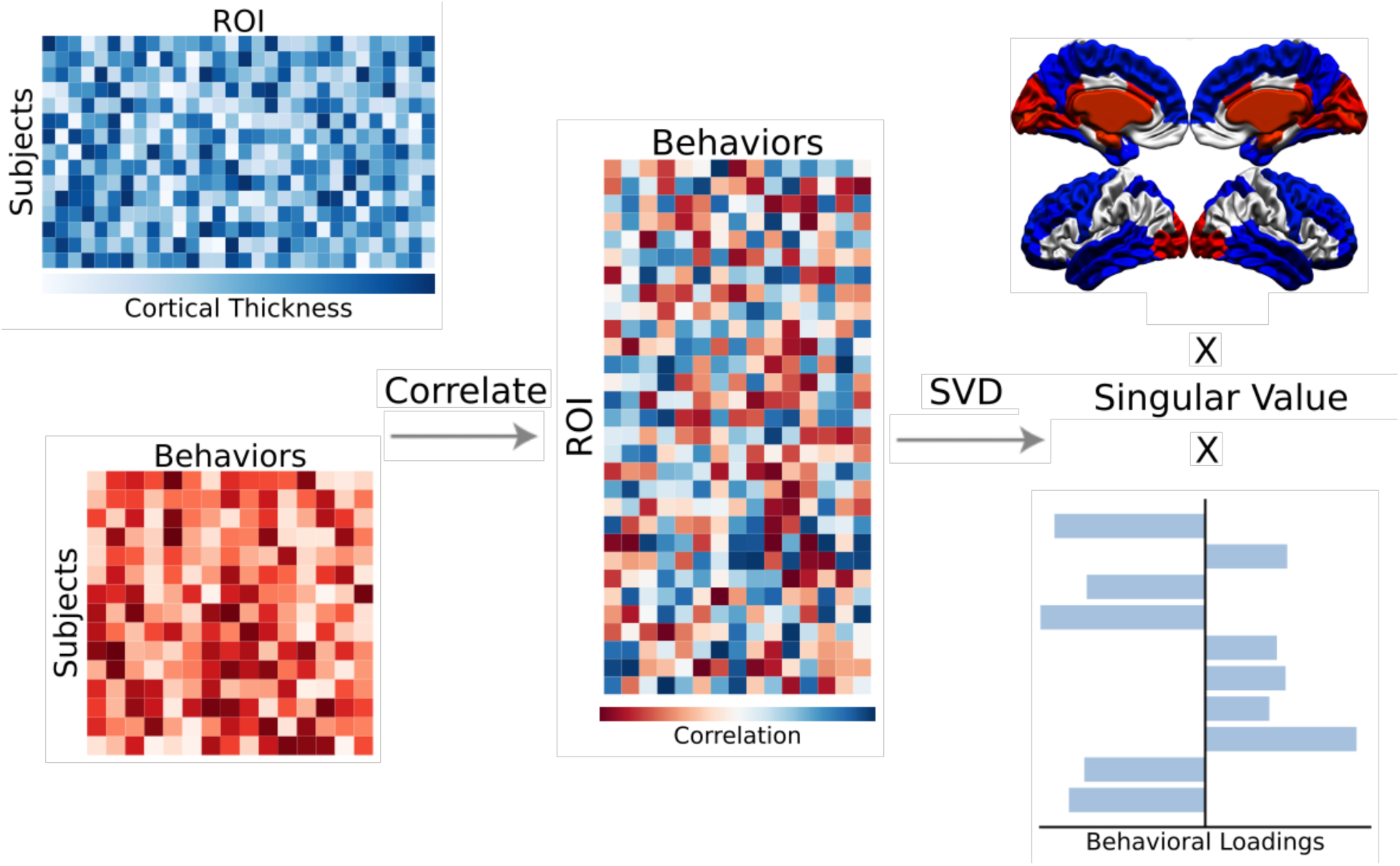
Workflow for Partial Least Squares (PLS) Analysis. PLS was used to identify patterns of covariance between regional cortical thickness measures and expression of lifestyle risk factor behavior for each participant in the analysis.

The statistical significance of each LV was assessed through permutation testing (5,000 permutations), where the rows of the brain matrix are randomly shuffled and the LV is recomputed (Krishnan et al., 2011). The reliability of the contribution of each brain and behavior variable was calculated through bootstrap resampling (5,000 bootstraps). This process takes samples, with replacement, from the participants, and reassesses the contribution of each variable to the LV to ensure that the pattern is not solely dependent on which participants are included in the sample. This reliability is measured as a bootstrap ratio (BSR), which is calculated by multiplying the weight of the variable in the LV by the singular value of that LV and dividing by the standard error (McIntosh & Lobaugh, 2004). A BSR of 1.95 was used, which is analogous to a p value of 0.05.

Additionally, we assessed the split-half stability of the LVs using a split-half resampling method, in which, for each PLS analysis, we split the sample into two halves, matching for age, sex, and menopause status where appropriate, and ran PLS on each half. For each resulting PLS model, we calculated the Pearson correlation between the cortical thickness and risk factor loadings separately (e.g. correlation between risk factor loadings for first half and second half). We took the absolute value of each correlation coefficient to account for arbitrary sign flips and repeated this process for 200 iterations. Finally, we used a metric proposed by McIntosh (2022) and Nakua et al. (2024) to determine if the resulting Pearson correlation coefficient distributions were significant, suggesting consistent relationships regardless of characteristics of participants. For each distribution, we performed a Z-test (i.e., mean divided by standard deviation of distribution) and a *Z*-test magnitude greater than 1.96, which is associated with a *p* value < .05, indicating that the distribution significantly differed from zero. Results are provided in the **Supplementary Figures.**

##### 2.5.2.1. Sex Differences Analysis

We performed PLS using the entire sample to elucidate common signatures of relationships between vertex-wise cortical thickness and lifestyle factors across sexes. All regression analyses were performed using R/4.2.2. Age and sex were not included in our input matrices and the impact was examined post-hoc using linear regression models to test the interaction of sex and age on PLS-derived brain and behavior scores (Eq 3).

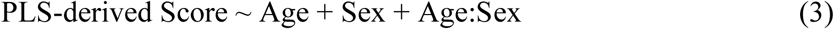

##### 2.5.2.2 Sex-Specific Analysis and the impact of menopause status

To explore sex-specific relationships, we performed sex-specific PLS, separating males and females and using identical lifestyle risk factors. This approach aimed to highlight any unique patterns within each sex that might not be observable in a combined sample. The entire male cohort was analyzed, but only female participants with a reported menopause status were included (n = 14,428) (Costantino et al, 2023). To examine the relationship of these sex-specific patterns to the aging process, we ran linear models using PLS-derived scores as the dependent variable and age as the independent variable (Eq 4).

##### 2.5.2.2 Sex-Specific Analysis and the impact of menopause status

To explore sex-specific relationships, we performed sex-specific PLS, separating males and females and using identical lifestyle risk factors. This approach aimed to highlight any unique patterns within each sex that might not be observable in a combined sample. The entire male cohort was analyzed, but only female participants with a reported menopause status were included (n = 14,428) (Costantino et al, 2023). To examine the relationship of these sex-specific patterns to the aging process, we ran linear models using PLS-derived scores as the dependent variable and age as the independent variable (Eq 4).

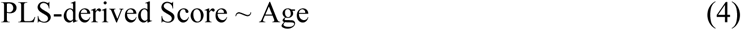

In females, to determine the effects of menopause status, we ran linear models to determine if there were interactions between menopause status and age on the PLS-derived scores (Eq 5).

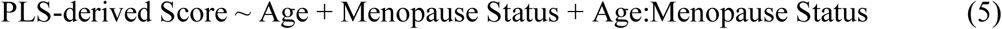

Additionally, to account for sex-specific factors, we repeated the female-specific PLS with the addition of binary menopause, surgical menopause, and natural menopause variables described above.

##### 2.5.2.3. Sex-Specific, Age-Matched Analysis

As age is the most significant risk factor for the development and progression of dementia, as well as for the onset of menopause, it is challenging to disentangle the effects of chronological aging from those of reproductive aging. Both processes contribute to neuroanatomical and cognitive changes but may do so through distinct mechanisms. For a more focused exploration of reproductive aging, rather than chronological aging, the menopause groups were age-matched using the nearest neighbors algorithm with the MatchIt package in R 4.1.2. An additional group of males was age-matched to the final female cohort. We performed PLS on the female group alone and with the male age-matched group included. In both analyses, an ANOVA was conducted to determine group differences in brain and risk factor pattern scores in any significant LVs found.

## 3. Results

### 3.1 Group Differences

A significant interaction between age and sex was observed over widespread regions of the cortex (**Figure 2A**), demonstrating age-related differences in cortical thickness that varied significantly by sex (p(FDR) < 0.05). More pronounced age-related cortical thinning was found in females compared to males in the frontal and temporal lobes with the effect sizes (standardized beta coefficients) ranging from 0.023 to 0.116.

**Figure 2:**
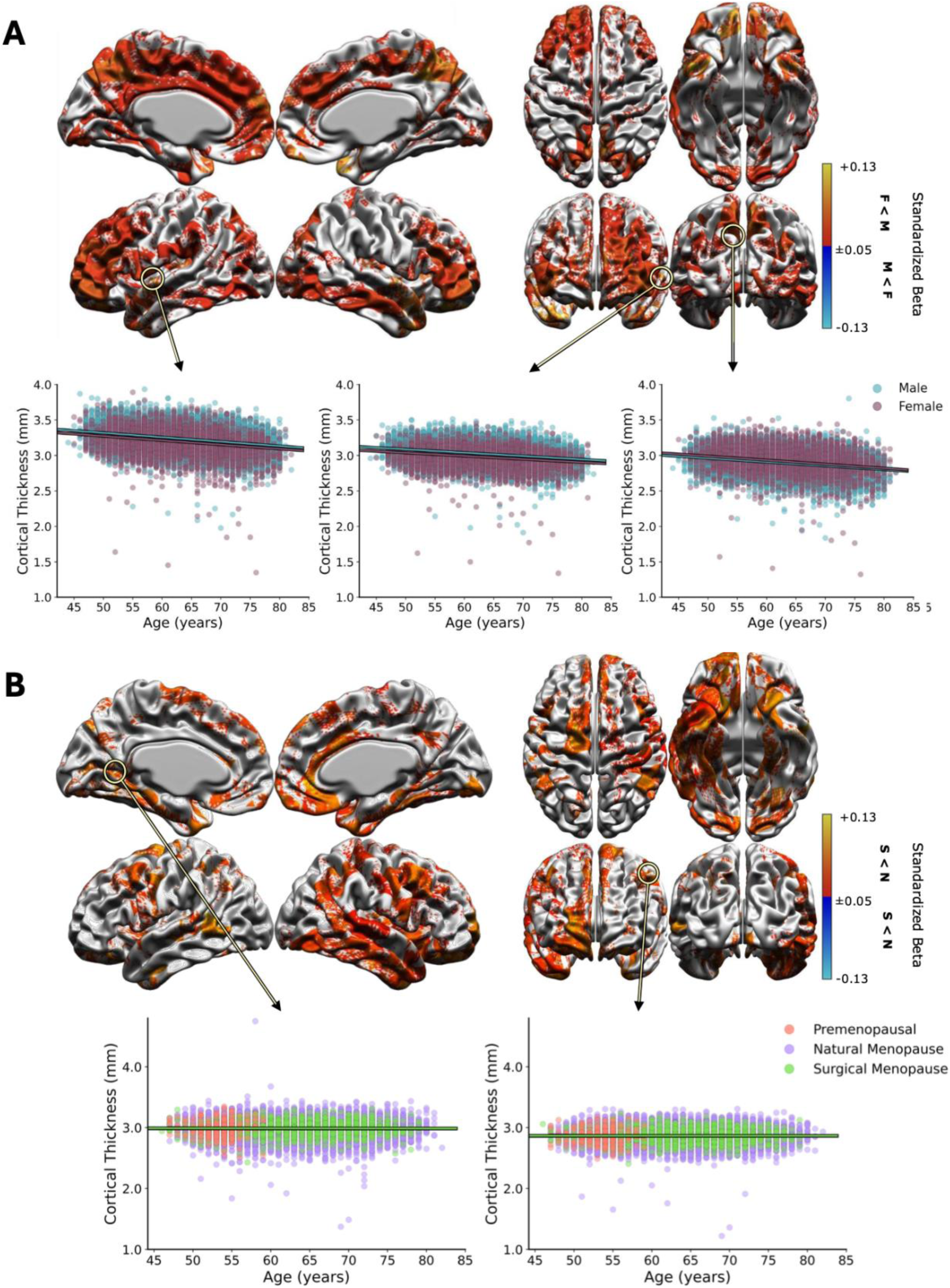
Results of linear regression analysis for group differences. Brain heatmaps showing significant age by female sex interaction, (**A**) and age by menopause status (**B**), colors indicate standardized beta coefficients. Plots of peak vertices are shown in the second row, stratified by sex and menopause status, respectively. Female sex*age at 5% FDR and Menopause status*age at 20%FDR (**F**: Female; **M:** Males**; N:** Natural menopause **S:** Surgical menopause).

For the female sample, a significant interaction (FDR-corrected p < 0.05) between age and menopause status was only observed between the surgical menopause and the natural menopause groups (**Figure 2B**), demonstrating thinner cortex in the surgical menopause group with age, relative to the natural menopause group with effect sizes ranging from 0.05 to 0.13. These differences were most widespread in the right hemisphere, covering the temporal, inferior parietal, and middle frontal areas. In the left hemisphere, effects were sparser, but more pronounced in the caudal middle frontal area.

### 3.2 Partial Least Squares Analysis

For all PLS analyses, detailed analyses were restricted to the first two significant LVs or to those that accounted for over 95 percent of the covariance if more than two were found.

**Figures 3A-B** display the brain and behavioral patterns for the corresponding latent variables in the whole sample and sex-specific analyses containing only the lifestyle risk factors. **Figures 3C-D** display the results for their respective linear regression models. In processing these analyses, we aimed to determine similarities and differences between the whole sample analysis and the sex-specific analyses, as well as between the sex-specific analyses themselves. There are multiple manifestations of differences that were observed: 1) factors that were significant in the whole sample analysis, but were not significant in either the male-or female-specific analyses; 2) factors that emerged only in the sex-specific analyses and not in whole sample analysis; and 3) factors that appeared in exclusively in the analysis of one sex, but were absent in the other sex or the whole sample.

**Figure 3:**
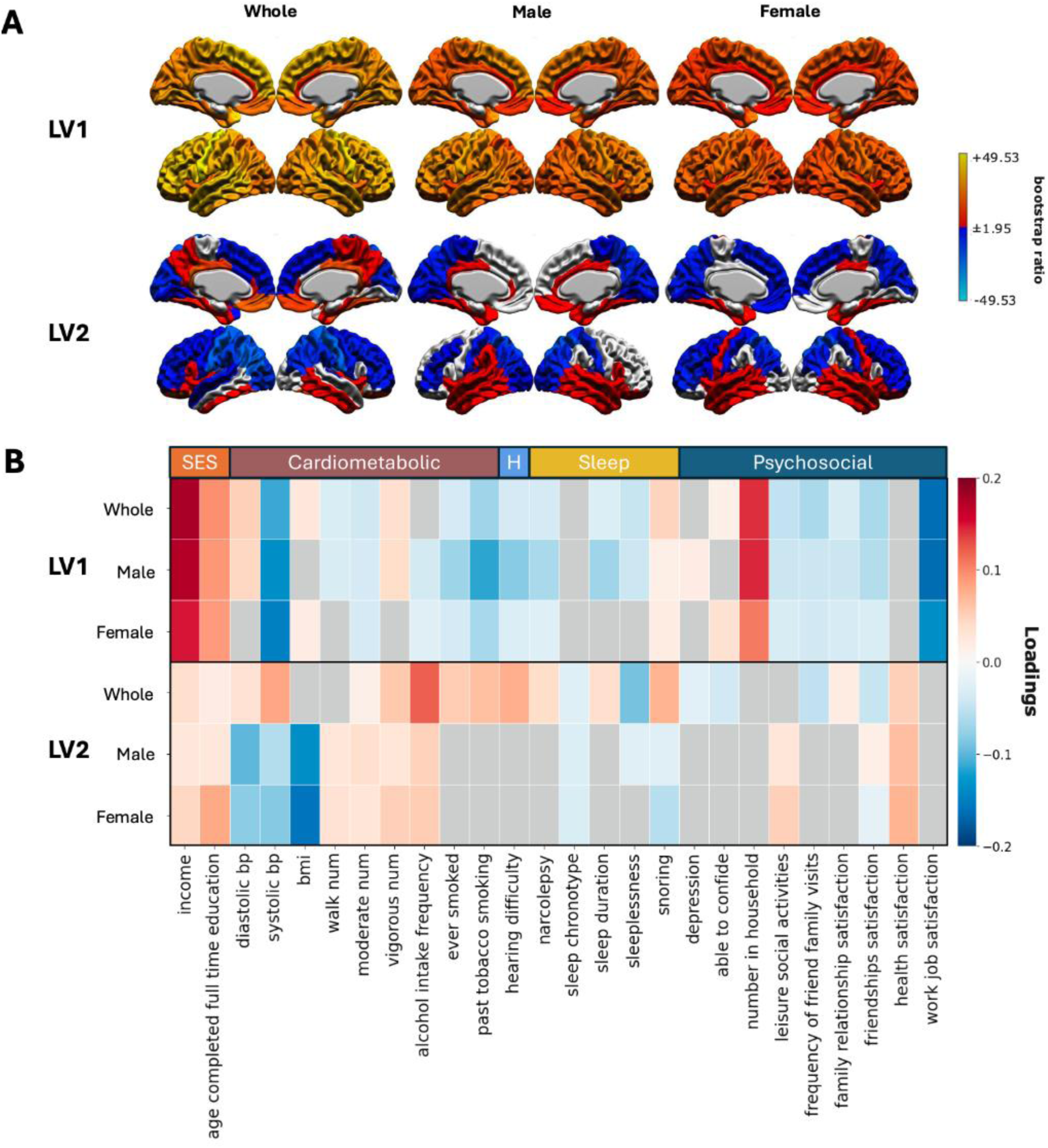
Results of whole sample, male-specific, and female-specific analyses. Morphometric **(A)** and risk factor **(B)** patterns for LV1 (top) and LV2 (bottom). Warm colors on morphometric maps indicate regions that vary positively with the LV, and cold colors indicate regions that vary negatively. These values are thresholded at ±1.95 bootstrap ratio (95% confidence interval). The x-axis on the risk factor plot indicates individual risk factors and color indicates the direction of the association with LV. Warm indicates positive association and cooler colors indicate negative loadings. Grey boxes indicate non-significant factors, as assessed by bootstrapping. (**H:** Hearing Difficulty).

**Figure 4:**
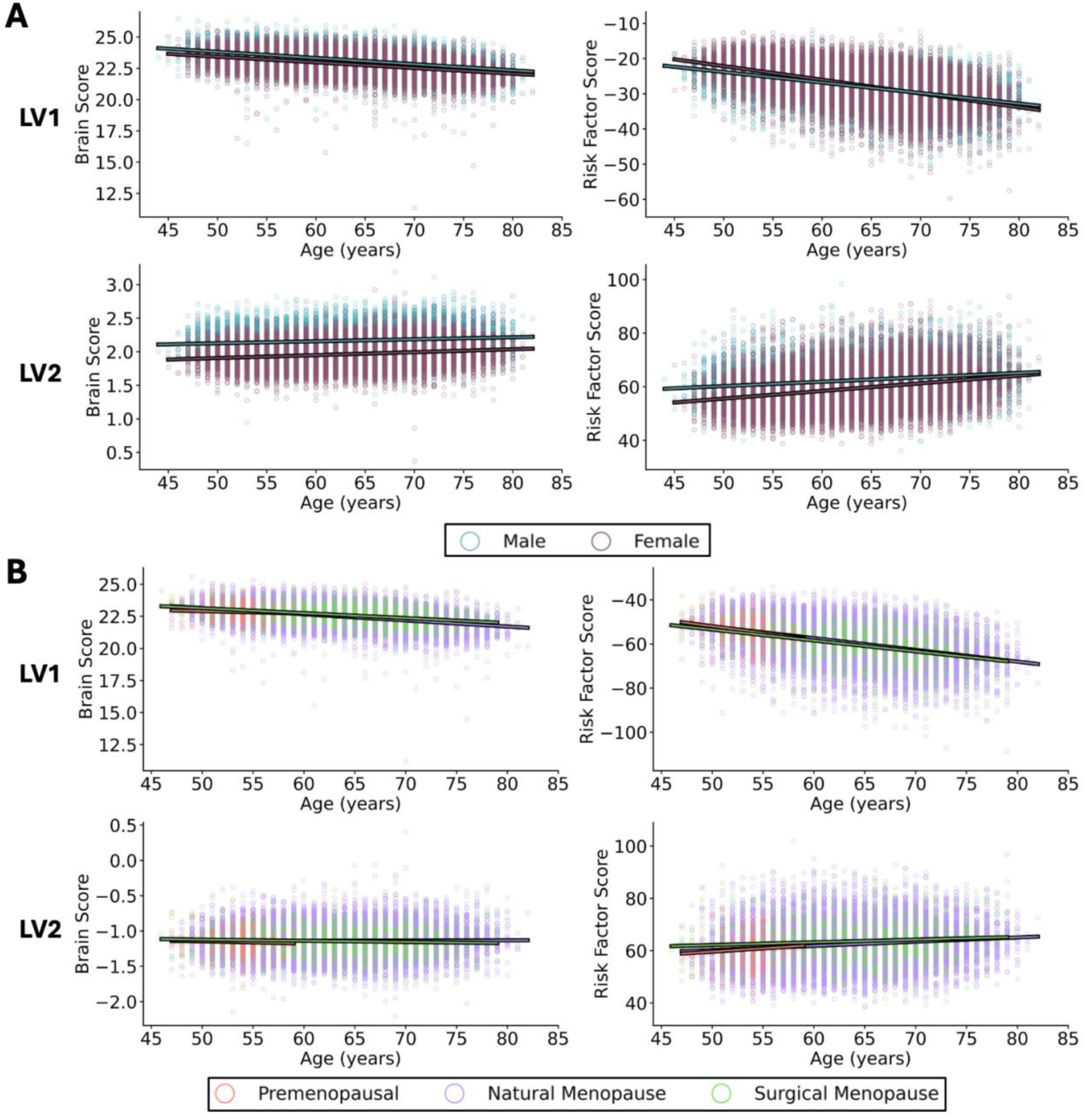
Interaction analyses of PLS-derived LV scores. Linear models used to test sex differences in the whole sample analysis (**A**) and menopause type differences in the female-specific analysis (**B**) for each LV-derived brain and risk factor scores for LV1 (top row) and LV2 (bottom row).

#### 3.2.1 Whole Sample Analysis

The PLS analysis yielded 7 significant (p < 0.05) LVs (**Supplementary Figure 1)** and the cortical thickness and risk factor loadings for the first 5 LVs were reproducible in split-half analysis (**Supplementary Figure 5)**. LV1 explained 91.71% (p < 0.001) of the covariance and was associated with significant contributions of factors associated with preserved brain health, such as higher socioeconomic class (e.g., increased educational attainment and higher income), more frequent vigorous exercise, and no history of smoking. In contrast, maladaptive sleep-related factors (e.g., shorter sleep duration and snoring) and lower indicators of social interaction (e.g., fewer leisure and social activities) and subjective well-being (e.g., lower satisfaction in friendships, family relationships and work) were also significant contributors. Other significant contributions include higher diastolic blood pressure, lower systolic blood pressure, higher BMI, lower frequency of walking and moderate exercise, fewer experiences of sleeplessness, and feeling able to confide in others. The covarying morphometry pattern involved positive associations with cortical thickness across the whole brain, strongest in the precentral gyrus, superior frontal gyrus, superior and transverse temporal regions.

LV2 explained 4.36% (p < 0.001) of the covariance and demonstrated significant contributions more associated with greater dementia risk, including worsened cardiometabolic health (e.g., higher diastolic and systolic blood pressure), increased substance use (e.g., increased alcohol consumption and a history of smoking) and hearing difficulty. Significant sleep-related contributions were mixed, with contributions related to both better sleep quality (e.g., longer sleep duration, earlier sleep chronotype, and lack of sleeplessness) and disturbed sleep patterns (e.g., snoring and narcolepsy). Social contact and satisfaction contributions were also mixed, with positive greater satisfaction in health and family relationships but feeling less able to confide in others, fewer friend and family visits, and lower friendship satisfaction. Contributions of higher socioeconomic status (e.g., increased educational attainment and higher income), exercise, and snoring were similar to those observed in LV1. Other significant contributions were increased frequency of vigorous and moderate exercise and an absence of depression. The covarying morphometric pattern included negative associations with cortical thickness across most of the frontal, parietal, and occipital regions, strongest in the postcentral gyrus regions in both hemispheres and the inferior parietal region in the left hemisphere and the superior parietal region in the right hemisphere. Both hemispheres had strong positive associations in the posterior cingulate, isthmus cingulate, and medial and lateral orbitofrontal regions.

The regression analysis revealed significant (p < 0.05) main effects of age and sex and a significant interaction between them on brain and behavior scores in both LV1 and LV2. For LV1, a negative main effect of age was observed for both brain (β = −0.424, p < 0.001) and behavior scores (β = −0.421, p < 0.001) on cortical thickness score, suggesting these patterns are more strongly associated with younger individuals. Sex showed opposing effects on brain and behavior scores, with a negative main effect on brain scores (β = −0.392, p < 0.001), suggesting stronger expression of the LV1 morphometric pattern in males, and a positive main effect on behavior scores (β = 0.091, p < 0.001), suggesting a stronger expression of the LV2 pattern in females. The interaction of age and sex also differed by domain. For brain scores, the interaction was positive (β = 0.340, p = 0.0008), indicating that with increasing age, females exhibited a more pronounced decrease in brain scores. In contrast, the interaction was negative for behavior scores (β = - 0.128, p < 0.001), suggesting males showed a rapid decrease in behavior scores for this pattern.

For LV2, a positive main effect of age was observed for brain scores (β = 0.104, p < 0.001) and behavior scores (β = 0.175, p < 0.001) on cortical thickness score, suggesting these patterns are more strongly associated with older individuals. The opposite was observed for sex, with negative main effects observed for both brain scores (β = −0.965, p < 0.001) and behavior scores (β = −0.445, p < 0.001), suggesting stronger expression of both patterns in males. The interaction of age and sex was positive for both brain scores (β = 0.490, p < 0.001) and behavior scores (β = - 0.130, p < 0.001), indicating, with increasing age, females showed a more pronounced increase in brain scores and behavior scores for this pattern.

#### 3.2.2 Sex-Specific Analysis

##### 3.2.2.1 Male-Specific Analysis

In males, the PLS analysis yielded 4 significant (p < 0.05) LVs (**Supplementary Figure 2**) and the cortical thickness and risk factor loadings for the first 3 LVs were reproducible in split-half analysis (**Supplementary Figure 5)**. LV1 accounted for 95.12% of the covariance (p < 0.05), with its significant risk factors and morphometric patterns closely resembling those identified in the whole sample analysis. Contributions from socioeconomic class, exercise, blood pressure, subjective well-being, and social enrichment remained consistent with the whole sample findings. However, the influence of snoring was reduced, while smoking history, hearing difficulty, and sleep disturbances showed stronger contributions. Depression and reduced alcohol intake frequency emerged as significant factors, whereas BMI and feeling able to confide were no longer significant. The associated morphometric pattern largely overlapped with that of the whole sample, showing positive associations with cortical thickness across the brain but with reduced loadings in the anterior cingulate regions.

LV2 accounted for 2.66% of the covariance (p < 0.05) and was linked to a risk factor profile markedly distinct from that of the whole sample analysis, showing a stronger association with preserved brain health. Significant contributions included greater educational attainment, higher income, better cardiometabolic health (e.g., lowered diastolic and systolic blood pressure, lower BMI, and more frequent exercise), better sleep (e.g., less frequent sleeplessness, no snoring), and greater social interaction and satisfaction (e.g., more leisure and social activities, greater satisfaction with friendships and health). No significant associations were observed related to any smoking-related risk factors, though alcohol intake frequency was positively associated with this pattern. There is overlap with regions identified in the whole-sample analysis, particularly in the middle frontal, parietal, and occipital regions, but many of the effects are dampened. Some regions exhibit a reversal in the direction of cortical thickness changes when analyzed specifically in males (e.g., cortical thickness increase in precuneus, temporal, and occipital regions), and other areas no longer have significant associations with the pattern (e.g., anterior cingulate, prefrontal gyrus, precentral gyrus, rostral middle frontal and superior frontal areas).

The results of the regression analysis found significant negative effects of age on brain scores in LV1(β = −0.440, p < 0.001) but not in LV2 (β = −0.0898, p =0.284). However, age was a significant predictor of behavior scores in both LV1 (β = −0.4191, p < 0.001) and LV2 (β = - 0.026, p = 0.00182), showing a decline with increasing age and stronger associations with younger individuals.

##### 3.2.2.2 Female-Specific Analysis

In females, the PLS analysis with only lifestyle risk variables yielded 5 significant (p < 0.05) LVs (**Supplementary Figure 3)** and the cortical thickness and risk factor loadings for the first 5 LVs were reproducible (**Supplementary Figure 5)**. LV1 explained 91.89% (p < 0.05) of the covariance and also demonstrated similar behavioral contributions as LV1 in the whole sample analysis. Contributions from socioeconomic class, smoking history, exercise, hearing difficulty, and social enrichment remained consistent with the whole sample findings, but associations with sleep disturbances (e.g., sleep duration and sleeplessness) are not observed. As seen in the male analysis, lowered alcohol intake frequency emerged as a significant contributor, and BMI and feeling able to confide are retained from the whole sample analysis, which did not occur in the male analysis. There was no significant contribution of diastolic blood pressure, observed before, but lowered systolic blood pressure remained significant. Additionally, a significant contribution of less frequent moderate exercise was found, but there was no significant contribution of walking or vigorous exercise, which were found in the whole sample and male analysis. The covarying morphometry pattern involved the cortical thickness increases across all regions, overlapping with the whole sample and male analysis, with dampening in cingulate areas.

LV2 explained 4.73% (p < 0.05) of the covariance and demonstrated a similar risk factor pattern to LV2 of the male analysis, though no association with sleeplessness is observed and the contribution of friendship satisfaction became negative. Many of the significant risk factor contributions are stronger in females, apart from lowered diastolic blood pressure and increased moderate exercise. The covarying morphometric pattern was also similar to that of LV2 in the male analysis, with retained associations with decreased cortical thickness in superior parietal areas and widespread associations with increased cortical thickness in temporal regions. Associations with decreased cortical thickness in frontal regions are retained from the whole sample analysis, but associations with occipital, cingulate, paracentral, and supramarginal regions are not observed.

The results of the regression analysis found significant negative effects of age for brain scores in LV1(β = −0.039, p = 0.008) and LV2 (β = −0.006, p =0.008), as well as for behavior scores in LV1 (β = −0.406, p =0.008), and LV2 (β = −0.136, p < 0.001), showing a decline with increasing age and stronger associations with younger individuals.

Our menopause status and age analysis (**Figure 3B)** found a significant main effect of surgical menopause on behavior scores for LV1 and LV2. In LV1, surgical menopause (β = - 0.11177, p = 0.000753) and the interaction of age and surgical menopause (β = −0.1448, p < 0.001) had negative effects on brain scores relative to the postmenopausal reference group. In LV2, the effects of surgical menopause (β = 0.0863, p = 0.00923) and a significant interaction of age and surgical menopause (β = 0.0716, p = 0.0267) were positive, indicate that surgical menopause, both as a main effect and in its interaction with age, plays a significant role in predicting behavioral scores. In contrast, premenopausal status and its interaction with age do not significantly influence behavioral outcomes. No other significant effects were found for the other terms in the model, or for any terms in the brain model.

Repeated PLS analysis with the addition of the binary menopause status variables yielded 5 significant (p < 0.05) LVs (**Supplementary Figure 4A**) and the cortical thickness and risk factor loadings for the first 3 LVs were reproducible (**Supplementary Figure 5)**. LV1 and LV2 demonstrated identical morphometric and risk factor patterns to the previous analysis. LV1 explained 92.11% (p < 0.05) of the covariance and demonstrated additional significant positive contributions of surgical menopause, but negative contributions of being premenopausal or experiencing natural menopause. LV2 explained 4.47% (p < 0.05) of the covariance and showed no significant contributions of any of the menopause variables.

### 3.3 Age-Matched PLS Analysis

In the age- and menopause-status matched female cohort, the PLS analysis with the inclusion of age-matched males found one significant LV (**Supplementary Figure 4B**) accounting for 59.84% of the covariance. Additionally, the significant LV’s risk factor loadings were reproducible from split-half analysis, but the cortical thickness loadings were not (**Supplementary Figure 5**). This LV demonstrated significant contributions of less frequent walking, more people in the household, and more work satisfaction. The covarying morphometry pattern involved the cortical thickness increases across all regions. We did not find any statistically significant differences in either risk factor pattern scores (F(3) = 0.202, p = 0.895) or brain scores (F(3) = 1.024, p = 0.381) between the groups.

Repeated analysis with the exclusion of the male cohort yielded 2 significant (p < 0.05) LVs (**Supplementary Figure 4C**) and the cortical thickness and risk factor loadings were found to be reproducible for LV2, but not LV1 (**Supplementary Figure 5**). LV1 explained 60.89% (p < 0.05) of the covariance and demonstrated significant contributions of lower diastolic blood pressure, earlier sleep chronotype, and shorter sleep duration. The covarying morphometry pattern involved the cortical thickness increased across all regions. Neither brain scores (F(2) = 0.544, p - 0.581) or behavior scores (F(2) = 0.664, p - 0.515) were different between the menopausal groups. T LV2 explained 9.45% (p < 0.05) of the covariance and demonstrated significant contributions of depression and ability to confide. The covarying morphometric pattern included decreasing cortical thickness in frontal regions and increased cortical thickness in temporal regions. Neither brain scores (F(2) = 0.052, p = 0.95) or behavior scores (F(2) = 0.497, p = 0.609) were different between the menopausal groups.

## 4. Discussion

In this study, we aimed to investigate the impact of sex and menopause on the associations between known modifiable risk factors and cortical thickness in healthy aging in a large, well-powered cohort of middle-aged and older adults. Overall, we found effects of sex and surgical menopause on age-related decline across the frontal and temporal lobes. We computed regression models to determine the interaction between age and sex and age and menopause status, revealed regions with greater age-related cortical thinning in women relative to men, as well as in women who underwent surgical menopause relative to those who went through natural menopause, in regions associated with aging and cognition. We performed a series of PLS analyses to identify patterns of covariance between cortical thickness and dementia risk factor behaviors and assessed their relationships to the aging trajectory. We detected patterns that suggested sex-specific impacts of risk factors on brain anatomy, and a complex interaction between chronological aging and reproductive aging not fully disentangled by strict age-matching.

Our approach of a combined whole sample PLS and sex-specific PLS allowed us to unravel sex-specific relationships to healthy brain aging that may have been diluted or obscured in siloed analysis. For LV1 in all main analyses, widespread increased cortical thickness was significantly associated with risk factors profiles generally linked with the preservation of brain health in younger individuals, but alcohol intake frequency was only identified as a significant impact in the sex-specific analysis. Heavy alcohol consumption has been linked to the cortical thickness of the precentral gyrus, superior temporal gyrus, and the dorsolateral prefrontal cortex (Morris et al., 2019), regions strongly associated with LV1 for all analyses, but it is possible that greater variability in alcohol consumption or outcomes within one sex diluted the overall signal in the pooled analysis. Though men tend to drink more frequently than women (Wilsnack et al., 2009), studies that have found more severe neuroanatomical effects of alcohol substance use disorders in women, even with similar intake patterns (Seo et al., 2019; Srivastava et al., 2010; Verplaetse et al., 2021; White, 2020). Other factors showed impacts on one sex and not the other, such as BMI contributing significantly to this pattern in women and not in men. Body fat has been shown to have potential neuroprotective effects in women as a primary source of estrogen (Simpson, 2003), and similar measures of BMI in men and women may be indicative of differing distributions of adipose tissue post-puberty (Chang et al., 2018; Fried et al., 2015; Karastergiou et al., 2012). One study has suggested that measures of visceral adipose tissue, not BMI, serve as a better anthropometric measure of obesity in women, as it conferred a much stronger association with cardiometabolic and cardiovascular risk (Kammerlander et al., 2021). The inclusion of the binary menopause status factors in the female analysis did not change the observed risk factor patterns and showed contributions indicative of younger individuals in the sample, as both the premenopausal and surgical menopause group were younger than the postmenopausal group.

Additionally, many of the risk factor contributions retained from the whole sample analysis were stronger in males, relative to females, in ways that may be explained by sex-related differences in strength of impacts. Household size, which can be a reliable proxy measure of social isolation, contributed substantially to both patterns and has been associated with cortical thickness in the frontal pole, temporal cortex, and precuneus cortex (Cong et al., 2024), but men have been shown to both experience more social isolation (Umberson et al., 2022) and experience more mental deterioration in isolation than women (Dury et al., 2021).

In contrast, the opposing morphometric and risk factor patterns of LV2 differed between the whole sample analysis and the sex-specific analysis, particularly in their relationship to age. LV2 showed poorer cardiometabolic health related to reduced cortical thickness in widespread regions related to dorsal attention and executive systems but increased cortical thickness in regions associated with emotional and motivational systems, such as the anterior cingulate cortex and orbitofrontal cortex, a latent dimension observed in another study (Nicolaisen-Sobesky et al., 2024). This pattern was associated with older adults but was expressed more quickly with increasing age in women, perhaps due to greater vulnerability to the effects of cardiometabolic risk factors on cortical thickness seen in women (Kim et al., 2019; Zhernakova et al., 2022).

However, LV2 in the sex-specific analyses showed patterns of better cardiometabolic health and increased cortical thickness in temporal regions for both sexes, but reduction in parietal and occipital regions in women and frontal and parietal regions in men. In both sexes, the brain pattern showed no significant relationship to age, but the risk factor patterns showed a significant but subtle relationship to age in males and a similar age and menopause status-surgical versus natural menopause-interaction in females. These findings concur with evidence that cardiometabolic influences on neural health emerge more gradually in men, but also that women who underwent surgical menopause, who have been shown to experience more prevalent cardiometabolic disturbances (Farahmand et al., 2015), may show greater vulnerability of these areas to accelerated aging due to hormonal insufficiency.

Our results from the PLS with strict age-matching replicated previous findings showing that, when isolating the impacts of menopause at midlife, we find no differences between the menopause groups or their male-counterparts in either risk factor patterns or behavioral patterns, which aligns with similar analyses from our group (Costantino et al., 2023; Wezel et al., 2024). For the females, this aligns with the findings of the PLS analyses with menopause-variables included, which did not change any of the observed risk factor or morphometric patterns found in their absence. For LV1, the menopause-related variable contributions were indicative of younger individuals in the sample, as both the premenopausal and surgical menopause groups were younger than the postmenopausal group, which aligns with greater association of that LV with younger individuals. Moreover, none of the menopause-related variables contributed significantly to any of the remaining LVs in that analysis. However, we did find significantly faster age-related cortical thinning in women who underwent surgical menopause in the vertex-wise models of cortical thickness, which may indicate that higher resolution neuroanatomical measures may be necessary to detect menopause-related differences.

### 4.1. Limitations

First, a limitation of this study is the reliance on cross-sectional data from the UK Biobank, which precludes direct inferences about the mechanistic or causal nature of the latent associations identified and limits the ability to make definitive claims about their relationship to aging. Second, it has been shown that there are biases in self-report accuracy in biobank-scale research (Schoeler et al., 2023). A large majority of the risk factor data was collected in the form of questionnaires, and and self-reported levels of factors like physical activity can be both underreported or over reported due to recall bias in older adults (Falck et al., 2020) or responses aimed at socially desirable responses(Althubaiti, 2016). Additionally, this study was limited to the characterization of each lifestyle risk factor available from the lifestyle information collected by the UK Biobank. While extensive, it was found that it limited the ability to fully capture each risk factor or the dimensions of each risk factor. For example, depression is considered a risk factor for dementia, and our analysis was limited to a third-party derived variable of a probable major depression as a binary input variable - this “diagnosis” category was defined using items related to lifetime experience of depressive symptoms (Smith et al., 2013). Moreover, many other variables were captured as categorical variables that were dummy coded to discrete integers, which may impact the ability of the PLS analysis to meaningfully explore the covariance patterns of the risk factor variables as would be possible with more continuous variables. This process could be improved by the addition of a more extensive set of behavioral variables that capture more aspects of each behavior and potential impacts on cortical thickness. Additionally, repeated analysis with male and female cohorts better matched across behavioral variables would allow us to explore the results of the PLS analysis more accurately in the context of sex differences.

## Data and Code Availability

The data supporting this study’s findings are available through the UK Biobank application procedure (https://www.ukbiobank.ac.uk/enable-your-research/register); scripts are available from the authors upon request.

## Funding

This work was undertaken thanks in part to support from the Canadian Institutes of Health Research (CIHR), the Natural Sciences and Engineering Research Council of Canada (NSERC), le Fonds de recherche du Québec – Santé (FRQS), and the Canada First Research Excellence Fund (CFREF), awarded through the Healthy Brains, Healthy Lives (HBHL) initiative at McGill University, with additional salary support from FRQS and HBHL.

## Author Contributions

**AM**: Conceptualization, Formal Analysis, Writing. **MC**: Conceptualization, Data Curation, Quality Control. **OP**: Data Curation. **GD**: Data Curation, Analysis Support. **MMC**: Conceptualization, Supervision, Funding Acquisition

## Declaration of Competing Interests

The authors have no competing interests to declare.

## Data Availability

All data supporting this study's findings are available through the UK Biobank application procedure (https://www.ukbiobank.ac.uk/enable-your-research/register)

## Notes

### Competing Interest Statement

The authors have declared no competing interest.

### Author Declarations

UK Biobank has obtained Research Tissue Bank (RTB) approval from the Research Ethics Committee (REC) (reference: 16/NW/0274). This research has been conducted under Application No. 45551.

## References

Ad-Dab’bagh, Y., Lyttelton, O., Muehlboeck, J.-S., Lepage, C., Einarson, D., Mok, K., Ivanov, O., Vincent, R., Lerch, J., Fombonne, E., & Evans, A. (2006, June). The CIVET image-processing environment: A fully automated comprehensive pipeline for anatomical neuroimaging research. Proceedings of the 12th Annual Meeting of the Organization for Human Brain Mapping.

Althubaiti, A. (2016). Information bias in health research: Definition, pitfalls, and adjustment methods. Journal of Multidisciplinary Healthcare, 9, 211–217. 10.2147/JMDH.S104807

Alzheimer’s Association. (2023). 2023 Alzheimer’s disease facts and figures. Alzheimer’s & Dementia, 19(4), 1598–1695. 10.1002/alz.13016

Anttila, T., Helkala, E.-L., Viitanen, M., Kåreholt, I., Fratiglioni, L., Winblad, B., Soininen, H., Tuomilehto, J., Nissinen, A., & Kivipelto, M. (2004). Alcohol drinking in middle age and subsequent risk of mild cognitive impairment and dementia in old age: A prospective population based study. BMJ: British Medical Journal, 329(7465), 539. 10.1136/bmj.38181.418958.BE

Azad, N. A., Al Bugami, M., & Loy-English, I. (2007). Gender differences in dementia risk factors. Gender Medicine, 4(2), 120–129. 10.1016/S1550-8579(07)80026-X

Barth, C., & de Lange, A.-M. G. (2020). Towards an understanding of women’s brain aging: The immunology of pregnancy and menopause. Frontiers in Neuroendocrinology, 58, 100850. 10.1016/j.yfrne.2020.100850

Barth, C., Galea, L. A., Jacobs, E. G., Lee, B. H., Westlye, L. T., & Lange, A.-M. G. de. (2024). Menopausal hormone therapy and the female brain: Leveraging neuroimaging and prescription registry data from the UK Biobank cohort. eLife, 13. 10.7554/eLife.99538.1

Bedford, S. A., Park, M. T. M., Devenyi, G. A., Tullo, S., Germann, J., Patel, R., Anagnostou, E., Baron-Cohen, S., Bullmore, E. T., Chura, L. R., Craig, M. C., Ecker, C., Floris, D. L., Holt, R. J., Lenroot, R., Lerch, J. P., Lombardo, M. V., Murphy, D. G. M., Raznahan, A.,…Chakravarty, M. M. (2020). Large-scale analyses of the relationship between sex, age and intelligence quotient heterogeneity and cortical morphometry in autism spectrum disorder. Molecular Psychiatry, 25(3), 614–628. 10.1038/s41380-019-0420-6

Benito-León, J., Bermejo-Pareja, F., Vega, S., & Louis, E. D. (2009). Total daily sleep duration and the risk of dementia: A prospective population-based study. European Journal of Neurology, 16(9), 990–997. 10.1111/j.1468-1331.2009.02618.x

Benjamini, Y., & Hochberg, Y. (1995). Controlling the False Discovery Rate: A Practical and Powerful Approach to Multiple Testing. Journal of the Royal Statistical Society. Series B (Methodological), 57(1), 289–300.

Chang, E., Varghese, M., & Singer, K. (2018). Gender and Sex Differences in Adipose Tissue. Current Diabetes Reports, 18(9), 69. 10.1007/s11892-018-1031-3

Conde, D. M., Verdade, R. C., Valadares, A. L. R., Mella, L. F. B., Pedro, A. O., & Costa-Paiva, L. (2021). Menopause and cognitive impairment: A narrative review of current knowledge. World Journal of Psychiatry, 11(8), 412–428. 10.5498/wjp.v11.i8.412

Cong, C.-H., Li, P.-L., Qiao, Y., Li, Y.-N., Yang, J.-T., Zhao, L., Zhu, X.-R., Tian, S., Cao, S.-S., Liu, J.-R., & Su, J.-J. (2024). Association between household size and risk of incident dementia in the UK Biobank study. Scientific Reports, 14(1), 11026. 10.1038/s41598-024-61102-6

Costantino, M., Pigeau, G., Parent, O., Ziolkowski, J., Devenyi, G. A., Gervais, N. J., & Chakravarty, M. M. (2023). Menopause, Brain Anatomy, Cognition and Alzheimer’s Disease. eLife, 12. 10.7554/eLife.91038.1

Crestol, A., de Lange, A.-M. G., Schindler, L., Subramaniapillai, S., Nerland, S., Oppenheimer, H., Westlye, L. T., Andreassen, O. A., Agartz, I., Tamnes, C. K., & Barth, C. (2024). Linking menopause-related factors, history of depression, APOE ε4, and proxies of biological aging in the UK biobank cohort. Hormones and Behavior, 164, 105596. 10.1016/j.yhbeh.2024.105596

Desikan, R. S., Ségonne, F., Fischl, B., Quinn, B. T., Dickerson, B. C., Blacker, D., Buckner, R. L., Dale, A. M., Maguire, R. P., Hyman, B. T., & others. (2006). An automated labeling system for subdividing the human cerebral cortex on MRI scans into gyral based regions of interest. Neuroimage, 31(3), 968–980.

Dury, S., Stas, L., Switsers, L., Duppen, D., Domènech-Abella, J., Dierckx, E., & Donder, L. D. (2021). Gender-related differences in the relationship between social and activity participation and health and subjective well-being in later life. Social Science & Medicine, 270, 113668. 10.1016/j.socscimed.2020.113668

Falck, R. S., Hsu, C. L., Best, J. R., Li, L. C., Egbert, A. R., & Liu-Ambrose, T. (2020). Not Just for Joints: The Associations of Moderate-to-Vigorous Physical Activity and Sedentary Behavior with Brain Cortical Thickness. Medicine & Science in Sports & Exercise, 52(10), 2217. 10.1249/MSS.0000000000002374

Farahmand, M., Ramezani Tehrani, F., Bahri Khomami, M., Noroozzadeh, M., & Azizi, F. (2015). Surgical menopause versus natural menopause and cardio-metabolic disturbances: A 12-year population-based cohort study. Journal of Endocrinological Investigation, 38(7), 761–767. 10.1007/s40618-015-0253-3

Fessel, J. (2023). The several ways to authentically cure Alzheimer’s dementia. Ageing Research Reviews, 92, 102093. 10.1016/j.arr.2023.102093

Fried, S. K., Lee, M.-J., & Karastergiou, K. (2015). Shaping fat distribution: New insights into the molecular determinants of depot-and sex-dependent adipose biology. Obesity, 23(7), 1345–1352. 10.1002/oby.21133

Gong, J., Harris, K., Lipnicki, D. M., Castro-Costa, E., Lima-Costa, M. F., Diniz, B. S., Xiao, S., Lipton, R. B., Katz, M. J., Wang, C., Preux, P.-M., Guerchet, M., Gbessemehlan, A., Ritchie, K., Ancelin, M.-L., Skoog, I., Najar, J., Sterner, T. R., Scarmeas, N.,…Consortium (COSMIC), for the C. S. of M. in an I. (2023). Sex differences in dementia risk and risk factors: Individual-participant data analysis using 21 cohorts across six continents from the COSMIC consortium. Alzheimer’s & Dementia, 19(8), 3365–3378. 10.1002/alz.12962

Government of Canada, S. C. (2022, April 27). A portrait of Canada’s growing population aged 85 and older from the 2021 Census. https://www12.statcan.gc.ca/census-recensement/2021/as-sa/98-200-X/2021004/98-200-X2021004-eng.cfm

Grodstein, F., Leurgans, S. E., Capuano, A. W., Schneider, J. A., & Bennett, D. A. (2023). Trends in Postmortem Neurodegenerative and Cerebrovascular Neuropathologies Over 25 Years. JAMA Neurology, 80(4), 370–376. 10.1001/jamaneurol.2022.5416

Hunt, J. F. V., Buckingham, W., Kim, A. J., Oh, J., Vogt, N. M., Jonaitis, E. M., Hunt, T. K., Zuelsdorff, M., Powell, R., Norton, D., Rissman, R. A., Asthana, S., Okonkwo, O. C., Johnson, S. C., Kind, A. J. H., & Bendlin, B. B. (2020). Association of Neighborhood-Level Disadvantage With Cerebral and Hippocampal Volume. JAMA Neurology, 77(4), 451–460. 10.1001/jamaneurol.2019.4501

Huo, N., Vemuri, P., Graff-Radford, J., Syrjanen, J., Machulda, M., Knopman, D. S., Jack, C. R., Petersen, R., & Mielke, M. M. (2022). Sex Differences in the Association Between Midlife Cardiovascular Conditions or Risk Factors With Midlife Cognitive Decline. Neurology, 98(6), e623–e632. 10.1212/WNL.0000000000013174

Kammerlander, A. A., Lyass, A., Mahoney, T. F., Massaro, J. M., Long, M. T., Vasan, R. S., & Hoffmann, U. (2021). Sex Differences in the Associations of Visceral Adipose Tissue and Cardiometabolic and Cardiovascular Disease Risk: The Framingham Heart Study. Journal of the American Heart Association: Cardiovascular and Cerebrovascular Disease, 10(11), e019968. 10.1161/JAHA.120.019968

Karastergiou, K., Smith, S. R., Greenberg, A. S., & Fried, S. K. (2012). Sex differences in human adipose tissues – the biology of pear shape. Biology of Sex Differences, 3(1), 13. 10.1186/2042-6410-3-13

Kim, S. E., Lee, J. S., Woo, S., Kim, S., Kim, H. J., Park, S., Lee, B. I., Park, J., Kim, Y., Jang, H., Kim, S. J., Cho, S. H., Lee, B., Lockhart, S. N., Na, D. L., & Seo, S. W. (2019). Sex-specific relationship of cardiometabolic syndrome with lower cortical thickness. Neurology, 93(11), e1045–e1057. 10.1212/WNL.0000000000008084

Kivimäki, M., Batty, G. D., Pentti, J., Shipley, M. J., Sipilä, P. N., Nyberg, S. T., Suominen, S. B., Oksanen, T., Stenholm, S., Virtanen, M., Marmot, M. G., Singh-Manoux, A., Brunner, E. J., Lindbohm, J. V., Ferrie, J. E., & Vahtera, J. (2020). Association between socioeconomic status and the development of mental and physical health conditions in adulthood: A multi-cohort study. The Lancet Public Health, 5(3), e140–e149. 10.1016/S2468-2667(19)30248-8

Klein, A., & Tourville, J. (2012). 101 Labeled Brain Images and a Consistent Human Cortical Labeling Protocol. Frontiers in Neuroscience, 6. 10.3389/fnins.2012.00171

Krishnan, A., Williams, L. J., McIntosh, A. R., & Abdi, H. (2011). Partial Least Squares (PLS) methods for neuroimaging: A tutorial and review. NeuroImage, 56(2), 455–475. 10.1016/j.neuroimage.2010.07.034

Liao, H., Cheng, J., Pan, D., Deng, Z., Liu, Y., Jiang, J., Cai, J., He, B., Lei, M., Li, H., Li, Y., Xu, Y., & Tang, Y. (2023). Association of earlier age at menopause with risk of incident dementia, brain structural indices and the potential mediators: A prospective community-based cohort study. eClinicalMedicine, 60. 10.1016/j.eclinm.2023.102033

Livingston, G., Huntley, J., Sommerlad, A., Ames, D., Ballard, C., Banerjee, S., Brayne, C., Burns, A., Cohen-Mansfield, J., Cooper, C., Costafreda, S. G., Dias, A., Fox, N., Gitlin, L. N., Howard, R., Kales, H. C., Kivimäki, M., Larson, E. B., Ogunniyi, A.,…Mukadam, N. (2020). Dementia prevention, intervention, and care: 2020 report of the Lancet Commission. The Lancet, 396(10248), 413–446. 10.1016/S0140-6736(20)30367-6

Lutsey, P. L., Misialek, J. R., Mosley, T. H., Gottesman, R. F., Punjabi, N. M., Shahar, E., MacLehose, R., Ogilvie, R. P., Knopman, D., & Alonso, A. (2018). Sleep characteristics and risk of dementia and Alzheimer’s disease: The Atherosclerosis Risk in Communities Study. Alzheimer’s & Dementia, 14(2), 157–166. 10.1016/j.jalz.2017.06.2269

McIntosh, A. R. (2022). Comparison of Canonical Correlation and Partial Least Squares analyses of simulated and empirical data (arXiv:2107.06867). arXiv. 10.48550/arXiv.2107.06867

McIntosh, A. R., & Lobaugh, N. J. (2004). Partial least squares analysis of neuroimaging data: Applications and advances. Mathematics in Brain Imaging, 23, S250–S263. 10.1016/j.neuroimage.2004.07.020

McIntosh, A. R., & Mišić, B. (2013). Multivariate Statistical Analyses for Neuroimaging Data. Annual Review of Psychology, 64(Volume 64, 2013), 499–525. 10.1146/annurev-psych-113011-143804

Miller, K. L., Alfaro-Almagro, F., Bangerter, N. K., Thomas, D. L., Yacoub, E., Xu, J., Bartsch, A. J., Jbabdi, S., Sotiropoulos, S. N., Andersson, J. L. R., Griffanti, L., Douaud, G., Okell, T. W., Weale, P., Dragonu, I., Garratt, S., Hudson, S., Collins, R., Jenkinson, M.,…Smith, S. M. (2016). Multimodal population brain imaging in the UK Biobank prospective epidemiological study. Nature Neuroscience, 19(11), 1523–1536. 10.1038/nn.4393

Morris, V. L., Owens, M. M., Syan, S. K., Petker, T. D., Sweet, L. H., Oshri, A., MacKillop, J., & Amlung, M. (2019). Associations Between Drinking and Cortical Thickness in Younger Adult Drinkers: Findings From the Human Connectome Project. Alcoholism: Clinical and Experimental Research, 43(9), 1918–1927. 10.1111/acer.14147

Mosconi, L., Berti, V., Dyke, J., Schelbaum, E., Jett, S., Loughlin, L., Jang, G., Rahman, A., Hristov, H., Pahlajani, S., Andrews, R., Matthews, D., Etingin, O., Ganzer, C., de Leon, M., Isaacson, R., & Brinton, R. D. (2021). Menopause impacts human brain structure, connectivity, energy metabolism, and amyloid-beta deposition. Scientific Reports, 11(1), 10867. 10.1038/s41598-021-90084-y

Nakua, H., Yu, J.-C., Abdi, H., Hawco, C., Voineskos, A., Hill, S., Lai, M.-C., Wheeler, A. L., McIntosh, A. R., & Ameis, S. H. (2024). Comparing the stability and reproducibility of brain-behavior relationships found using canonical correlation analysis and partial least squares within the ABCD sample. Network Neuroscience, 8(2), 576–596. 10.1162/netn_a_00363

Nichols, E., Steinmetz, J. D., Vollset, S. E., Fukutaki, K., Chalek, J., Abd-Allah, F., Abdoli, A., Abualhasan, A., Abu-Gharbieh, E., Akram, T. T., Hamad, H. A., Alahdab, F., Alanezi, F. M., Alipour, V., Almustanyir, S., Amu, H., Ansari, I., Arabloo, J., Ashraf, T.,…Vos, T. (2022). Estimation of the global prevalence of dementia in 2019 and forecasted prevalence in 2050: An analysis for the Global Burden of Disease Study 2019. The Lancet Public Health, 7(2), e105–e125. 10.1016/S2468-2667(21)00249-8

Nicolaisen-Sobesky, E., Maleki Balajoo, S., Mahdipour, M., Mihalik, A., Hoffstaedter, F., Mourao-Miranda, J., Tahmasian, M., Eickhoff, S. B., & Genon, S. (2024). Cardiometabolic health, cortical thickness, and neurotransmitter systems: A large-scale multivariate study. bioRxiv, 2024.06.14.599066. 10.1101/2024.06.14.599066

Ramli, N. Z., Yahaya, M. F., Mohd Fahami, N. A., Abdul Manan, H., Singh, M., & Damanhuri, H. A. (2023). Brain volumetric changes in menopausal women and its association with cognitive function: A structured review. Frontiers in Aging Neuroscience, 15, 1158001. 10.3389/fnagi.2023.1158001

Rocca, W. A., Grossardt, B. R., & Shuster, L. T. (2014). Oophorectomy, estrogen, and dementia: A 2014 update. Molecular and Cellular Endocrinology, 389(1), 7–12. 10.1016/j.mce.2014.01.020

Schoeler, T., Pingault, J.-B., & Kutalik, Z. (2023). Self-report inaccuracy in the UK Biobank: Impact on inference and interplay with selective participation (p. 2023.10.06.23296652). medRxiv. 10.1101/2023.10.06.23296652

Seo, S., Beck, A., Matthis, C., Genauck, A., Banaschewski, T., Bokde, A. L. W., Bromberg, U., Büchel, C., Quinlan, E. B., Flor, H., Frouin, V., Garavan, H., Gowland, P., Ittermann, B., Martinot, J.-L., Paillère Martinot, M.-L., Nees, F., Papadopoulos Orfanos, D., Poustka, L.,…Obermayer, K. (2019). Risk profiles for heavy drinking in adolescence: Differential effects of gender. Addiction Biology, 24(4), 787–801. 10.1111/adb.12636

Simpson, E. R. (2003). Sources of estrogen and their importance. The Journal of Steroid Biochemistry and Molecular Biology, 86(3), 225–230. 10.1016/S0960-0760(03)00360-1

Sindi, S., Kåreholt, I., Johansson, L., Skoog, J., Sjöberg, L., Wang, H.-X., Johansson, B., Fratiglioni, L., Soininen, H., Solomon, A., Skoog, I., & Kivipelto, M. (2018). Sleep disturbances and dementia risk: A multicenter study. Alzheimer’s & Dementia, 14(10), 1235–1242. 10.1016/j.jalz.2018.05.012

Smith, D. J., Nicholl, B. I., Cullen, B., Martin, D., Ul-Haq, Z., Evans, J., Gill, J. M. R., Roberts, B., Gallacher, J., Mackay, D., Hotopf, M., Deary, I., Craddock, N., & Pell, J. P. (2013). Prevalence and Characteristics of Probable Major Depression and Bipolar Disorder within UK Biobank: Cross-Sectional Study of 172,751 Participants. PLOS ONE, 8(11), e75362. 10.1371/journal.pone.0075362

Srivastava, V., Buzas, B., Momenan, R., Oroszi, G., Pulay, A. J., Enoch, M.-A., Hommer, D. W., & Goldman, D. (2010). Association of SOD2, a Mitochondrial Antioxidant Enzyme, with Gray Matter Volume Shrinkage in Alcoholics. Neuropsychopharmacology, 35(5), 1120– 1128. 10.1038/npp.2009.217

Strandberg, A. Y., Trygg, T., Pitkälä, K. H., & Strandberg, T. E. (2018). Alcohol consumption in midlife and old age and risk of frailty: Alcohol paradox in a 30-year follow-up study. Age and Ageing, 47(2), 248–254. 10.1093/ageing/afx165

Subramaniapillai, S., Schindler, L. S., Redmond, P., Bastin, M. E., Wardlaw, J. M., Valdés Hernández, M., Maniega, S. M., Aribisala, B., Westlye, L. T., Coath, W., Groves, J., Cash, D. M., Barnes, J., James, S.-N., Sudre, C. H., Barkhof, F., Richards, M., Corley, J., Russ, T. C.,…de Lange, A.-M. G. (2024). Sex-Dependent Effects of Cardiometabolic Health and APOE4 on Brain Age. Neurology, 103(6), e209744. 10.1212/WNL.0000000000209744

Subramaniapillai, S., Suri, S., Barth, C., Maximov, I. I., Voldsbekk, I., van der Meer, D., Gurholt, T. P., Beck, D., Draganski, B., Andreassen, O. A., Ebmeier, K. P., Westlye, L. T., & de Lange, A.-M. G. (2022). Sex-and age-specific associations between cardiometabolic risk and white matter brain age in the UK Biobank cohort. Human Brain Mapping, 43(12), 3759–3774. 10.1002/hbm.25882

Than, S., Moran, C., Collyer, T. A., Beare, R. J., Lane, E. M., Vincent, A. J., Wang, W., Callisaya, M. L., Thomson, R., Phan, T. G., Fornito, A., & Srikanth, V. K. (2022). Associations of Sex, Age, and Cardiometabolic Risk Profiles With Brain Structure and Cognition: A UK Biobank Latent Class Analysis. Neurology, 99(17), e1853–e1865. 10.1212/WNL.0000000000201028

Umberson, D., Lin, Z., & Cha, H. (2022). Gender and Social Isolation across the Life Course. Journal of Health and Social Behavior, 63(3), 319–335. 10.1177/00221465221109634

Verplaetse, T. L., Cosgrove, K. P., Tanabe, J., & McKee, S. A. (2021). Sex/gender differences in brain function and structure in alcohol use: A narrative review of neuroimaging findings over the last 10 years. Journal of Neuroscience Research, 99(1), 309–323. 10.1002/jnr.24625

Wezel, D., Parent, O., Costantino, M., Sifi, L., Pigeau, G., Gervais, N. J., McQuarrie, A., Maranzano, J., Devenyi, G. A., Dadar, M., & Chakravarty, M. M. (2024). Untangling age and menopausal status reveals no effect of menopause on white matter hyperintensity volume (p. 2024.10.28.24316270). medRxiv. 10.1101/2024.10.28.24316270

White, A. (2020). Gender Differences in the Epidemiology of Alcohol Use and Related Harms in the United States. Alcohol Research: Current Reviews, 40(2), 01. 10.35946/arcr.v40.2.01

Wilsnack, R. W., & Wilsnack, S. C. (2013). Gender and alcohol: Consumption and consequences. In Alcohol: Science, policy, and public health. (pp. 153–160). Oxford University Press. 10.1093/acprof:oso/9780199655786.003.0017

Wilsnack, R. W., Wilsnack, S. C., Kristjanson, A. F., Vogeltanz-Holm, N. D., & Gmel, G. (2009). Gender and alcohol consumption: Patterns from the multinational GENACIS project. Addiction, 104(9), 1487–1500. 10.1111/j.1360-0443.2009.02696.x

Xiong, L. Y., Wood Alexander, M., Wong, Y. Y., Wu, C.-Y., Ruthirakuhan, M., Edwards, J. D., Lanctôt, K. L., Black, S. E., Rabin, J. S., Cogo-Moreira, H., & Swardfager, W. (2024). >Latent profiles of modifiable dementia risk factors in later midlife: Relationships with incident dementia, cognition, and neuroimaging outcomes. Molecular Psychiatry, 1–11. 10.1038/s41380-024-02685-4

Zeighami, Y., Fereshtehnejad, S.-M., Dadar, M., Collins, D. L., Postuma, R. B., Mišić, B., & Dagher, A. (2019). A clinical-anatomical signature of Parkinson’s disease identified with partial least squares and magnetic resonance imaging. NeuroImage, 190, 69–78. 10.1016/j.neuroimage.2017.12.050

Zhernakova, D. V., Sinha, T., Andreu-Sánchez, S., Prins, J. R., Kurilshikov, A., Balder, J.-W., Sanna, S., Franke, L., Kuivenhoven, J. A., Zhernakova, A., & Fu, J. (2022). Age-dependent sex differences in cardiometabolic risk factors. Nature Cardiovascular Research, 1(9), 844–854. 10.1038/s44161-022-00131-8

