## Supplemental Material for "Investigating the impact of sex and reproductive aging on latent signatures of modifiable dementia risk factors"

### S. Supplementary Materials

### S1. Supplementary Tables

| Non-cancer Illness Coding | Meaning | N |
|---|---|---|
| 1081 | Stroke | 230 |
| 1082 | Transient ischaemic attack (tia) | 160 |
| 1083 | Subdural haemorrhage/haematoma | 7 |
| 1086 | Subarachnoid haemorrhage | 7 |
| 1240 | Neurological injury/trauma | 4 |
| 1243 | Psychological/psychiatric problem | 1 |
| 1244 | Infection of nervous system | 0 |
| 1245 | Brain abscess/intracranial abscess | 2 |
| 1246 | Encephalitis | 12 |
| 1247 | Meningitis | 92 |
| 1258 | Chronic/degenerative neurological problem | 2 |
| 1259 | Motor neurone disease | 3 |
| 1261 | Multiple sclerosis | 79 |
| 1262 | Parkinson's disease | 44 |
| 1263 | Dementia/Alzheimers/cognitive impairment | 8 |
| 1264 | Epilepsy | 116 |
| 1266 | Head injury | 31 |
| 1408 | Alcohol dependency | 19 |
| 1409 | Opioid dependency | 0 |
| 1410 | Other substance abuse/dependency | 0 |
| 1434 | Other neurological problem | 17 |
| 1491 | Brain haemorrhage | 12 |
| 1583 | Ischaemic stroke | 10 |
| 1626 | Fracture skull / head | 31 |

**Table S1:** Non-cancer illness codes used to exclude participants due to neurological, psychiatric, and substance-related diagnoses.

| Final Variable Name | Data Field |
|---|---|
| Age | 21003 |
| Sex | 31 |
| Alcohol drinker status | 20117 |
| Alcohol intake frequency | 1558 |
| Diastolic blood pressure | 94, 4079 |
| Systolic blood pressure | 93, 4080 |
| Diabetes | 2443 |
| Age completed full time education | 845, 6138 |
| Hearing difficulty | 2247, 2257 |
| Hypertension | 2966, 6150 |
| Depression | 20123, 20124, 20125 |
| Body mass index (BMI) | 23104 |
| Waist-to-hip ratio | 48, 49 |
| Duration of moderate activity | 894 |
| Duration of walks | 874 |
| Duration of vigorous activity | 914 |
| Number of days/week walked 10+ minutes | 864 |
| Number of days/week of moderate physical activity 10+ minutes | 884 |
| Number of days/week of vigorous physical activity 10+ minutes | 904 |
| Average total household income before tax | 738 |
| Sleep duration | 1160 |
| Morning-evening person (chronotype) | 1180 |
| Sleeplessness/insomnia | 1200 |
| Snoring | 1210 |
| Daytime dozing/sleeping (narcolepsy) | 1220 |
| Smoking status | 20116 |
| Past tobacco smoking | 1249 |
| Pack years of smoking | 20161 |
| Able to confide | 2110 |
| Frequency of friend-family visits | 1031 |
| Number in household | 709 |
| Leisure-social activities | 6160 |
| Work-job satisfaction | 4537 |
| Health satisfaction | 4548 |
| Family relationship satisfaction | 4559 |
| Friendships satisfaction | 4570 |
| Medication for cholesterol, blood pressure or diabetes | 6177 |
| Ever smoked | 20160 |

**Table S2:** Variable field codes from the UK Biobank used to derive final analysis variables

| N | Female<br>17,503 | Male<br>14,208 | p-value |
|---|---|---|---|
| **Age (years)** | 63.1 (7.0) | 63.8 (7.6) | **< 0.001** |
| **Income** | | | **< 0.001** |
| Less than 18,000 | 2414 (13.8%) | 1319 (9.3%) | |
| 18,000 to 30,999 | 5237 (29.9%) | 3559 (25.0%) | |
| 31,000 to 51,999 | 5443 (31.1%) | 4428 (31.2%) | |
| 52,000 to 100,000 | 3441 (19.7%) | 3720 (26.2%) | |
| Greater than 100,000 | 968 (5.5%) | 1182 (8.3%) | |
| **Age completed full time education (years)** | 18.9 (2.5) | 19.1 (2.6) | **< 0.001** |
| **Diastolic Blood Pressure (mmHg)** | 77.0 (9.4) | 80.6 (9.3) | **< 0.001** |
| **Systolic Blood Pressure (mmHg)** | 135.2 (17.9) | 141.2 (16.2) | **< 0.001** |
| **Body Mass Index (BMI)** | 26.0 (4.6) | 27.0 (3.8) | **< 0.001** |
| **Frequency of walking** | 5.8 (1.7) | 5.6 (1.8) | **< 0.001** |
| **Frequency of moderate exercise** | 4.1 (2.2) | 3.9 (2.2) | **< 0.001** |
| **Frequency of vigorous exercise** | 1.8 (1.8) | 2.2 (1.9) | **< 0.001** |
| **Alcohol Intake Frequency** | | | **< 0.001** |
| Never | 1330 (7.6%) | 736 (5.2%) | |
| Special occasions only | 2351 (13.4%) | 950 (6.7%) | |
| One to three times a month | 2330 (13.3%) | 1339 (9.4%) | |
| Once or twice a week | 4765 (27.2%) | 3679 (25.9%) | |
| Three or four times a week | 4404 (25.2%) | 4566 (32.1%) | |
| Daily or almost daily | 2323 (13.3%) | 2938 (20.7%) | |
| **Ever Smoked** | | | **< 0.001** |
| No | 8838 (50.5%) | 6207 (43.7%) | |
| Yes | 8665 (49.5%) | 8001 (56.3%) | |
| **Past Tobacco Smoking** | | | **< 0.001** |
| I have never smoked | 8791 (50.2%) | 6185 (43.5%) | |
| Just tried once or twice | 2806 (16.0%) | 2333 (16.4%) | |
| Smoked occasionally | 2327 (13.3%) | 1724 (12.1%) | |
| Smoked on most or all days | 3579 (20.4%) | 3966 (27.9%) | |
| **Hearing Difficulty** | | | **< 0.001** |
| No | 10694 (61.1%) | 7079 (49.8%) | |
| Yes | 6809 (38.9%) | 7129 (50.2%) | |
| **Narcolepsy** | | | **< 0.001** |
| Never/rarely | 13966 (79.8%) | 10576 (74.4%) | |
| Sometimes | 3187 (18.2%) | 3263 (23.0%) | |
| Often | 350 (2.0%) | 369 (2.6%) | |
| All of the time | 0 (0%) | 0 (0%) | |
| **Sleep Chronotype** | | | 0.226 |
| Morning person | 11748 (67.1%) | 9444 (66.5%) | |
| Evening person | 5755 (32.9%) | 4764 (33.5%) | |
| **Sleep Duration** | 7.1 (1.0) | 7.2 (1.0) | |
| **Sleeplesness** | | | |
| Never/rarely | 2765 (15.8%) | 4076 (28.7%) | |
| Sometimes | 8481 (48.5%) | 6239 (43.9%) | |
| Usually | 6257 (35.7%) | 3893 (27.4%) | |
| **Snoring** | | | **< 0.001** |
| No | 12777 (73.0%) | 7999 (56.3%) | |
| Yes | 4726 (27.0%) | 6209 (43.7%) | |
| **Depression** | | | **< 0.001** |

| | | | |
|---|---|---|---|
| None | 16135 (92.2%) | 13437 (94.6%) | |
| Single episode of probably major depression | 351 (2.0%) | 196 (1.4%) | |
| Probable recurrent major depression (moderate) | 716 (4.1%) | 352 (2.5%) | |
| Probable recurrent major depression (severe) | 301 (1.7%) | 223 (1.6%) | |
| **Able to Confide** | | | 0.530 |
| Never or almost never | 1655 (9.5%) | 2450 (17.2%) | |
| Once every few months | 1116 (6.4%) | 945 (6.7%) | |
| About once a month | 1146 (6.5%) | 779 (5.5%) | |
| About once a week | 2325 (13.3%) | 1414 (10.0%) | |
| 2-4 times a week | 2149 (12.3%) | 1183 (8.3%) | |
| Almost daily | 9112 (52.1%) | 7437 (52.3%) | |
| **Number in Household** | 2.1 (1.0) | 2.2 (1.0) | 0.127 |
| **Leisure/Social Activities** | 1.3 (0.9) | 1.1 (0.9) | 0.407 |
| **Frequency of Friend/Family Visits** | | | 0.636 |
| No friends/family outside household | 16 (0.1%) | 17 (0.1%) | |
| Never or almost never | 132 (0.8%) | 250 (1.8%) | |
| Once every few months | 846 (4.8%) | 1099 (7.7%) | |
| About once a month | 1897 (10.8%) | 2198 (15.5%) | |
| About once a week | 5526 (31.6%) | 5110 (36.0%) | |
| 2-4 times a week | 6494 (37.1%) | 4367 (30.7%) | |
| Almost daily | 2592 (14.8%) | 1167 (8.2%) | |
| **Friendships Satisfaction** | | | 0.759 |
| Extremely unhappy | 9 (0.1%) | 25 (0.2%) | |
| Very unhappy | 42 (0.2%) | 48 (0.3%) | |
| Moderately unhappy | 329 (1.9%) | 452 (3.2%) | |
| Moderately happy | 4627 (26.4%) | 4879 (34.3%) | |
| Very happy | 9729 (55.6%) | 7306 (51.4%) | |
| Extremely happy | 2767 (15.8%) | 1498 (10.5%) | |
| **Health Satisfaction** | | | 0.467 |
| Extremely unhappy | 88 (0.5%) | 34 (0.2%) | |
| Very unhappy | 220 (1.3%) | 159 (1.1%) | |
| Moderately unhappy | 1183 (6.8%) | 828 (5.8%) | |
| Moderately happy | 7744 (44.2%) | 6327 (44.5%) | |
| Very happy | 7140 (40.8%) | 5908 (41.6%) | |
| Extremely happy | 1128 (6.4%) | 952 (6.7%) | |
| **Work/Job Satisfaction** | | | < **.001** |
| Extremely unhappy | 777 (4.4%) | 906 (6.4%) | |
| Very unhappy | 3470 (19.8%) | 3516 (24.7%) | |
| Moderately unhappy | 3564 (20.4%) | 3308 (23.3%) | |
| Moderately happy | 1153 (6.6%) | 572 (4.0%) | |
| Very happy | 803 (4.6%) | 139 (1.0%) | |
| Extremely happy | 222 (1.3%) | 69 (0.5%) | |
| I am not employed | 7514 (42.9%) | 5698 (40.1%) | |

**Table S3:** Sample demographics of the whole sample, separated by sex

| | | | | p-value | |
|---|---|---|---|---|---|
| | Premenopausal | Natural Menopause | Surgical Menopause | Pre vs Natural | Pre vs Surgical |
| N | 222 | 13,227 | 979 | | |
| **Age (years)** | 53.4 (2.4) | 63.4 (6.6) | 63.7 (6.9) | **< 0.001** | **< 0.001** |
| **Income** | | | | **< 0.001** | **< 0.001** |
| Less than 18,000 | 14 (6.3%) | 1810 (13.7%) | 127 (13.0%) | | |
| 18,000 to 30,999 | 28 (12.6%) | 4021 (30.4%) | 281 (28.7%) | | |
| 31,000 to 51,999 | 80 (36.0%) | 4116 (31.1%) | 364 (37.2%) | | |
| 52,000 to 100,000 | 72 (32.4%) | 2584 (19.5%) | 161 (16.4%) | | |
| Greater than 100,000 | 28 (12.6%) | 696 (5.3%) | 46 (4.7%) | | |
| **Age completed full time education (years)** | 19.3 (2.0) | 18.9 (2.5) | 18.4 (2.6) | **0.020** | **< 0.001** |
| **Diastolic Blood Pressure (mmHg)** | 76.2 (8.3) | 76.9 (9.4) | 77.5 (9.5) | 0.288 | 0.071 |
| **Systolic Blood Pressure (mmHg)** | 126.0 (14.7) | 135.2 (17.9) | 136.5 (18.1) | **< 0.001** | **< 0.001** |
| **Body Mass Index (BMI)** | 26.2 (4.6) | 25.9 (4.5) | 26.5 (4.4) | 0.305 | 0.116 |
| **Frequency of walking** | 5.6 (1.9) | 5.8 (1.6) | 5.7 (1.8) | 0.249 | 0.832 |
| **Frequency of moderate exercise** | 3.6 (2.2) | 4.1 (2.2) | 4.0 (2.2) | **< 0.001** | **0.013** |
| **Frequency of vigorous exercise** | 1.9 (1.9) | 1.9 (1.8) | 1.7 (1.8) | 0.650 | 0.183 |
| **Alcohol Intake Frequency** | | | | 0.080 | 0.251 |
| Never | 10 (4.5%) | 1002 (7.6%) | 86 (8.8%) | | |
| Special occasions only | 30 (13.5%) | 1698 (12.8%) | 156 (15.9%) | | |
| One to three times a month | 36 (16.2%) | 1739 (13.1%) | 140 (14.3%) | | |
| Once or twice a week | 72 (32.4%) | 3586 (27.1%) | 274 (28.0%) | | |
| Three or four times a week | 44 (19.8%) | 3434 (26.0%) | 198 (20.2%) | | |
| Daily or almost daily | 30 (13.5%) | 1768 (13.4%) | 125 (12.8%) | | |
| **Ever Smoked** | | | | 0.061 | 0.283 |
| No | 126 (56.8%) | 6638 (50.2%) | 514 (52.5%) | | |
| Yes | 96 (43.2%) | 6589 (49.8%) | 465 (47.5%) | | |
| **Past Tobacco Smoking** | | | | **0.036** | 0.056 |
| I have never smoked | 126 (56.8%) | 6607 (50.0%) | 514 (52.5%) | | |
| Just tried once or twice | 40 (18.0%) | 2141 (16.2%) | 135 (13.8%) | | |
| Smoked occasionally | 27 (12.2%) | 1781 (13.5%) | 138 (14.1%) | | |
| Smoked on most or all days | 29 (13.1%) | 2698 (20.4%) | 192 (19.6%) | | |
| **Hearing Difficulty** | | | | **< 0.001** | **< 0.001** |
| No | 163 (73.4%) | 8080 (61.1%) | 586 (59.9%) | | |
| Yes | 59 (26.6%) | 5147 (38.9%) | 393 (40.1%) | | |
| **Narcolepsy** | | | | 0.535 | 0.811 |
| Never/rarely | 183 (82.4%) | 10550 (79.8%) | 792 (80.9%) | | |
| Sometimes | 34 (15.3%) | 2408 (18.2%) | 167 (17.1%) | | |
| Often | 5 (2.3%) | 269 (2.0%) | 20 (2.0%) | | |
| All of the time | 0 (0%) | 0 (0%) | 0 (0%) | | |
| **Sleep Chronotype** | | | | 0.672 | 0.828 |
| Morning person | 146 (65.8%) | 8907 (67.3%) | 654 (66.8%) | | |
| Evening person | 76 (34.2%) | 4320 (32.7%) | 325 (33.2%) | | |
| **Sleep Duration** | 7.2 (1.0) | 7.1 (1.0) | 7.1 (1.2) | 0.331 | 0.347 |
| **Sleeplesness** | | | | **< 0.001** | **< 0.001** |
| Never/rarely | 69 (31.1%) | 2085 (15.8%) | 145 (14.8%) | | |
| Sometimes | 92 (41.4%) | 6403 (48.4%) | 461 (47.1%) | | |
| Usually | 61 (27.5%) | 4739 (35.8%) | 373 (38.1%) | | |
| **Snoring** | | | | 0.388 | 0.285 |
| No | 169 (76.1%) | 9697 (73.3%) | 708 (72.3%) | | |

| | | | | | |
|---|---|---|---|---|---|
| Yes | 53 (23.9%) | 3530 (26.7%) | 271 (27.7%) | | |
| **Depression** | | | | **0.0342** | 0.742 |
| None | 199 (89.6%) | 12210 (92.3%) | 876 (89.5%) | | |
| Single episode of probably major depression | 5 (2.3%) | 266 (2.0%) | 22 (2.2%) | | |
| Probable recurrent major depression (moderate) | 9 (4.1%) | 544 (4.1%) | 52 (5.3%) | | |
| Probable recurrent major depression (severe) | 9 (4.1%) | 207 (1.6%) | 29 (3.0%) | | |
| **Able to Confide** | | | | 0.759 | 0.291 |
| Never or almost never | 18 (8.1%) | 1210 (9.1%) | 116 (11.8%) | | |
| Once every few months | 18 (8.1%) | 849 (6.4%) | 51 (5.2%) | | |
| About once a month | 12 (5.4%) | 884 (6.7%) | 62 (6.3%) | | |
| About once a week | 26 (11.7%) | 1810 (13.7%) | 106 (10.8%) | | |
| 2-4 times a week | 29 (13.1%) | 1643 (12.4%) | 106 (10.8%) | | |
| Almost daily | 119 (53.6%) | 6831 (51.6%) | 538 (55.0%) | | |
| **Number in Household** | 2.8 (1.3) | 2.1 (1.0) | 2.0 (1.1) | **< 0.001** | **< 0.001** |
| **Leisure/Social Activities** | 1.1 (0.9) | 1.3 (0.9) | 1.2 (0.9) | **0.003** | **0.325** |
| **Frequency of Friend/Family Visits** | | | | **0.007** | **0.027** |
| No friends/family outside household | 1 (0.5%) | 13 (0.1%) | 2 (0.2%) | | |
| Never or almost never | 2 (0.9%) | 98 (0.7%) | 5 (0.5%) | | |
| Once every few months | 12 (5.4%) | 624 (4.7%) | 41 (4.2%) | | |
| About once a month | 36 (16.2%) | 1418 (10.7%) | 111 (11.3%) | | |
| About once a week | 77 (34.7%) | 4122 (31.2%) | 294 (30.0%) | | |
| 2-4 times a week | 76 (34.2%) | 4944 (37.4%) | 373 (38.1%) | | |
| Almost daily | 18 (8.1%) | 2008 (15.2%) | 153 (15.6%) | | |
| **Friendships Satisfaction** | | | | 0.207 | 0.537 |
| Extremely unhappy | 0 (0%) | 5 (0.0%) | 3 (0.3%) | | |
| Very unhappy | 0 (0%) | 28 (0.2%) | 2 (0.2%) | | |
| Moderately unhappy | 3 (1.4%) | 259 (2.0%) | 12 (1.2%) | | |
| Moderately happy | 71 (32.0%) | 3461 (26.2%) | 262 (26.8%) | | |
| Very happy | 107 (48.2%) | 7399 (55.9%) | 527 (53.8%) | | |
| Extremely happy | 41 (18.5%) | 2075 (15.7%) | 173 (17.7%) | | |
| **Health Satisfaction** | | | | **0.014** | **0.002** |
| Extremely unhappy | 2 (0.9%) | 53 (0.4%) | 10 (1.0%) | | |
| Very unhappy | 3 (1.4%) | 144 (1.1%) | 16 (1.6%) | | |
| Moderately unhappy | 20 (9.0%) | 809 (6.1%) | 83 (8.5%) | | |
| Moderately happy | 87 (39.2%) | 5776 (43.7%) | 464 (47.4%) | | |
| Very happy | 84 (37.8%) | 5555 (42.0%) | 362 (37.0%) | | |
| Extremely happy | 26 (11.7%) | 890 (6.7%) | 44 (4.5%) | | |
| **Work/Job Satisfaction** | | | | < **.001** | **< 0.001** |
| Extremely unhappy | 4 (1.8%) | 595 (4.5%) | 13 (1.3%) | | |
| Very unhappy | 15 (6.8%) | 2703 (20.4%) | 40 (4.1%) | | |
| Moderately unhappy | 15 (6.8%) | 2736 (20.7%) | 60 (6.1%) | | |
| Moderately happy | 79 (35.6%) | 782 (5.9%) | 178 (18.2%) | | |
| Very happy | 61 (27.5%) | 526 (4.0%) | 180 (18.4%) | | |
| Extremely happy | 24 (10.8%) | 134 (1.0%) | 47 (4.8%) | | |
| I am not employed | 24 (10.8%) | 5751 (43.5%) | 461 (47.1%) | | |

**Table S4:** Sample demographics of the female sample, separated by menopause status

| | | | | | p-value | | |
|---|---|---|---|---|---|---|---|
| | **Premenopausal** | **Natural Menopause** | **Surgical Menopause** | **Age-Matched Males** | **Pre vs Natural** | **Pre vs Surgical** | **Pre vs Males** |
| **N** | 222 | 445 | 223 | 890 | | | |
| **Age (years)** | 53.4 (2.4) | 53.8 (2.7) | 54.1 (3.0) | 53.8 (2.7) | 0.071 | **0.002** | **0.0485** |
| **Income** | | | | | 0.304 | 0.190 | **0.0129** |
| Less than 18,000 | 14 (6.3%) | 31 (7.0%) | 24 (10.8%) | 46 (5.2%) | | | |
| 18,000 to 30,999 | 28 (12.6%) | 71 (16.0%) | 37 (16.6%) | 108 (12.1%) | | | |
| 31,000 to 51,999 | 80 (36.0%) | 126 (28.3%) | 82 (36.8%) | 226 (25.4%) | | | |
| 52,000 to 100,000 | 72 (32.4%) | 163 (36.6%) | 58 (26.0%) | 361 (40.6%) | | | |
| Greater than 100,000 | 28 (12.6%) | 54 (12.1%) | 22 (9.9%) | 149 (16.7%) | | | |
| **Age completed full time education (years)** | 19.3 (2.0) | 19.1 (2.4) | 18.7 (2.6) | 19.2 (2.5) | 0.354 | **0.007** | 0.618 |
| **Diastolic Blood Pressure (mmHg)** | 76.2 (8.3) | 75.5 (9.0) | 78.0 (8.4) | 81.4 (9.5) | 0.280 | **0.026** | **< 0.001** |
| **Systolic Blood Pressure (mmHg)** | 126.0 (14.7) | 125.0 (14.2) | 127.7 (15.4) | 135.3 (14.0) | 0.622 | 0.265 | **< 0.001** |
| **Body Mass Index (BMI)** | 26.2 (4.6) | 25.7 (4.6) | 27.0 (5.2) | 27.2 (4.2) | 0.123 | 0.059 | **< 0.001** |
| **Frequency of walking** | 5.6 (1.9) | 5.6 (1.8) | 5.6 (1.9) | 5.5 (1.9) | 0.682 | 0.787 | 0.457 |
| **Frequency of moderate exercise** | 3.6 (2.2) | 3.8 (2.3) | 3.5 (2.3) | 3.6 (2.2) | 0.327 | 0.730 | 0.984 |
| **Frequency of vigorous exercise** | 1.9 (1.9) | 1.9 (1.8) | 1.4 (1.6) | 2.4 (1.9) | 0.703 | **0.016** | **< 0.001** |
| **Alcohol Intake Frequency** | | | | | 0.576 | **0.058** | **< 0.001** |
| Never | 10 (4.5%) | 26 (5.8%) | 20 (9.0%) | 59 (6.6%) | | | |
| Special occasions only | 30 (13.5%) | 63 (14.2%) | 43 (19.3%) | 55 (6.2%) | | | |
| One to three times a month | 36 (16.2%) | 64 (14.4%) | 39 (17.5%) | 101 (11.3%) | | | |
| Once or twice a week | 72 (32.4%) | 141 (31.7%) | 63 (28.3%) | 273 (30.7%) | | | |
| Three or four times a week | 44 (19.8%) | 107 (24.0%) | 42 (18.8%) | 266 (29.9%) | | | |
| Daily or almost daily | 30 (13.5%) | 44 (9.9%) | 16 (7.2%) | 136 (15.3%) | | | |
| **Ever Smoked** | | | | | 0.170 | 0.316 | **0.025** |
| No | 126 (56.8%) | 226 (50.8%) | 115 (51.6%) | 428 (48.1%) | | | |
| Yes | 96 (43.2%) | 219 (49.2%) | 108 (48.4%) | 462 (51.9%) | | | |
| **Past Tobacco Smoking** | | | | | 0.142 | 0.432 | **0.006** |
| I have never smoked | 126 (56.8%) | 226 (50.8%) | 115 (51.6%) | 425 (47.8%) | | | |
| Just tried once or twice | 40 (18.0%) | 68 (15.3%) | 36 (16.1%) | 137 (15.4%) | | | |
| Smoked occasionally | 27 (12.2%) | 68 (15.3%) | 36 (16.1%) | 124 (13.9%) | | | |
| Smoked on most or all days | 29 (13.1%) | 83 (18.7%) | 36 (16.1%) | 204 (22.9%) | | | |
| **Hearing Difficulty** | | | | | **0.009** | **0.012** | **0.005** |
| No | 163 (73.4%) | 280 (62.9%) | 138 (61.9%) | 561 (63.0%) | | | |
| Yes | 59 (26.6%) | 165 (37.1%) | 85 (38.1%) | 329 (37.0%) | | | |
| **Narcolepsy** | | | | | 0.722 | 0.540 | 0.983 |
| Never/rarely | 183 (82.4%) | 377 (84.7%) | 189 (84.8%) | 729 (81.9%) | | | |
| Sometimes | 34 (15.3%) | 58 (13.0%) | 27 (12.1%) | 140 (15.7%) | | | |
| Often | 5 (2.3%) | 10 (2.2%) | 7 (3.1%) | 21 (2.4%) | | | |
| All of the time | 0 (0%) | 0 (0%) | 0 (0%) | 0 (0%) | | | |
| **Sleep Chronotype** | | | | | 0.902 | 0.296 | 0.255 |
| Morning person | 146 (65.8%) | 289 (64.9%) | 135 (60.5%) | 546 (61.3%) | | | |
| Evening person | 76 (34.2%) | 156 (35.1%) | 88 (39.5%) | 344 (38.7%) | | | |
| **Sleep Duration** | 7.2 (1.0) | 7.0 (0.9) | 6.9 (1.3) | 7.0 (0.9) | | **0.009** | **0.001** |
| **Sleeplessness** | | | | | | **< 0.001** | 0.676 |
| Never/rarely | 69 (31.1%) | 76 (17.1%) | 34 (15.2%) | 290 (32.6%) | | | |
| Sometimes | 92 (41.4%) | 195 (43.8%) | 92 (41.3%) | 381 (42.8%) | | | |
| Usually | 61 (27.5%) | 174 (39.1%) | 97 (43.5%) | 219 (24.6%) | | | |
| **Snoring** | | | | | 0.911 | 0.294 | **<0.001** |
| No | 169 (76.1%) | 342 (76.9%) | 159 (71.3%) | 455 (51.1%) | | | |

| | | | | | | | |
|---|---|---|---|---|---|---|---|
| Yes | 53 (23.9%) | 103 (23.1%) | 64 (28.7%) | 435 (48.9%) | | | |
| **Depression** | | | | | 0.507 | 0.302 | **0.005** |
| None | 199 (89.6%) | 399 (89.7%) | 187 (83.9%) | 846 (95.1%) | | | |
| Single episode of probably major depression | 5 (2.3%) | 13 (2.9%) | 6 (2.7%) | 8 (0.9%) | | | |
| Probable recurrent major depression (moderate) | 9 (4.1%) | 23 (5.2%) | 17 (7.6%) | 26 (2.9%) | | | |
| Probable recurrent major depression (severe) | 9 (4.1%) | 10 (2.2%) | 13 (5.8%) | 10 (1.1%) | | | |
| **Able to Confide** | | | | | 0.530 | 0.228 | **0.026** |
| Never or almost never | 18 (8.1%) | 34 (7.6%) | 22 (9.9%) | 143 (16.1%) | | | |
| Once every few months | 18 (8.1%) | 21 (4.7%) | 8 (3.6%) | 55 (6.2%) | | | |
| About once a month | 12 (5.4%) | 24 (5.4%) | 12 (5.4%) | 51 (5.7%) | | | |
| About once a week | 26 (11.7%) | 60 (13.5%) | 18 (8.1%) | 81 (9.1%) | | | |
| 2-4 times a week | 29 (13.1%) | 50 (11.2%) | 27 (12.1%) | 82 (9.2%) | | | |
| Almost daily | 119 (53.6%) | 256 (57.5%) | 136 (61.0%) | 478 (53.7%) | | | |
| **Number in Household** | 2.8 (1.3) | 2.7 (1.2) | 2.5 (1.9) | 2.9 (1.3) | 0.127 | **< 0.001** | 0.915 |
| **Leisure/Social Activities** | 1.1 (0.9) | 1.0 (0.8) | 0.8 (0.8) | 1.0 (0.8) | 0.407 | **0.004** | 0.548 |
| **Frequency of Friend/Family Visits** | | | | | 0.636 | 0.772 | **0.001** |
| No friends/family outside household | 1 (0.5%) | 0 (0%) | 1 (0.4%) | 2 (0.2%) | | | |
| Never or almost never | 2 (0.9%) | 6 (1.3%) | 2 (0.9%) | 18 (2.0%) | | | |
| Once every few months | 12 (5.4%) | 23 (5.2%) | 18 (8.1%) | 96 (10.8%) | | | |
| About once a month | 36 (16.2%) | 63 (14.2%) | 35 (15.7%) | 163 (18.3%) | | | |
| About once a week | 77 (34.7%) | 176 (39.6%) | 68 (30.5%) | 365 (41.0%) | | | |
| 2-4 times a week | 76 (34.2%) | 137 (30.8%) | 73 (32.7%) | 198 (22.2%) | | | |
| Almost daily | 18 (8.1%) | 40 (9.0%) | 26 (11.7%) | 48 (5.4%) | | | |
| **Friendships Satisfaction** | | | | | 0.759 | 0.820 | **0.001** |
| Extremely unhappy | 0 (0%) | 0 (0%) | 1 (0.4%) | 3 (0.3%) | | | |
| Very unhappy | 0 (0%) | 1 (0.2%) | 0 (0%) | 10 (1.1%) | | | |
| Moderately unhappy | 3 (1.4%) | 11 (2.5%) | 4 (1.8%) | 51 (5.7%) | | | |
| Moderately happy | 71 (32.0%) | 142 (31.9%) | 71 (31.8%) | 314 (35.3%) | | | |
| Very happy | 107 (48.2%) | 219 (49.2%) | 111 (49.8%) | 419 (47.1%) | | | |
| Extremely happy | 41 (18.5%) | 72 (16.2%) | 36 (16.1%) | 93 (10.4%) | | | |
| **Health Satisfaction** | | | | | 0.467 | **0.003** | 0.189 |
| Extremely unhappy | 2 (0.9%) | 1 (0.2%) | 4 (1.8%) | 5 (0.6%) | | | |
| Very unhappy | 3 (1.4%) | 11 (2.5%) | 7 (3.1%) | 22 (2.5%) | | | |
| Moderately unhappy | 20 (9.0%) | 47 (10.6%) | 25 (11.2%) | 76 (8.5%) | | | |
| Moderately happy | 87 (39.2%) | 178 (40.0%) | 100 (44.8%) | 386 (43.4%) | | | |
| Very happy | 84 (37.8%) | 171 (38.4%) | 82 (36.8%) | 340 (38.2%) | | | |
| Extremely happy | 26 (11.7%) | 37 (8.3%) | 5 (2.2%) | 61 (6.9%) | | | |
| **Work/Job Satisfaction** | | | | | < **.001** | 0.228 | **< 0.001** |
| Extremely unhappy | 4 (1.8%) | 36 (8.1%) | 5 (2.2%) | 49 (5.5%) | | | |
| Very unhappy | 15 (6.8%) | 129 (29.0%) | 15 (6.7%) | 252 (28.3%) | | | |
| Moderately unhappy | 15 (6.8%) | 163 (36.6%) | 31 (13.9%) | 387 (43.5%) | | | |
| Moderately happy | 79 (35.6%) | 50 (11.2%) | 71 (31.8%) | 97 (10.9%) | | | |
| Very happy | 61 (27.5%) | 23 (5.2%) | 57 (25.6%) | 27 (3.0%) | | | |
| Extremely happy | 24 (10.8%) | 8 (1.8%) | 16 (7.2%) | 16 (1.8%) | | | |
| I am not employed | 24 (10.8%) | 36 (8.1%) | 28 (12.6%) | 62 (7.0%) | | | |

**Table S5:** Sample demographics of the age-matched menopause sample, separated by sex and menopause status.

### S2. Supplementary Figures

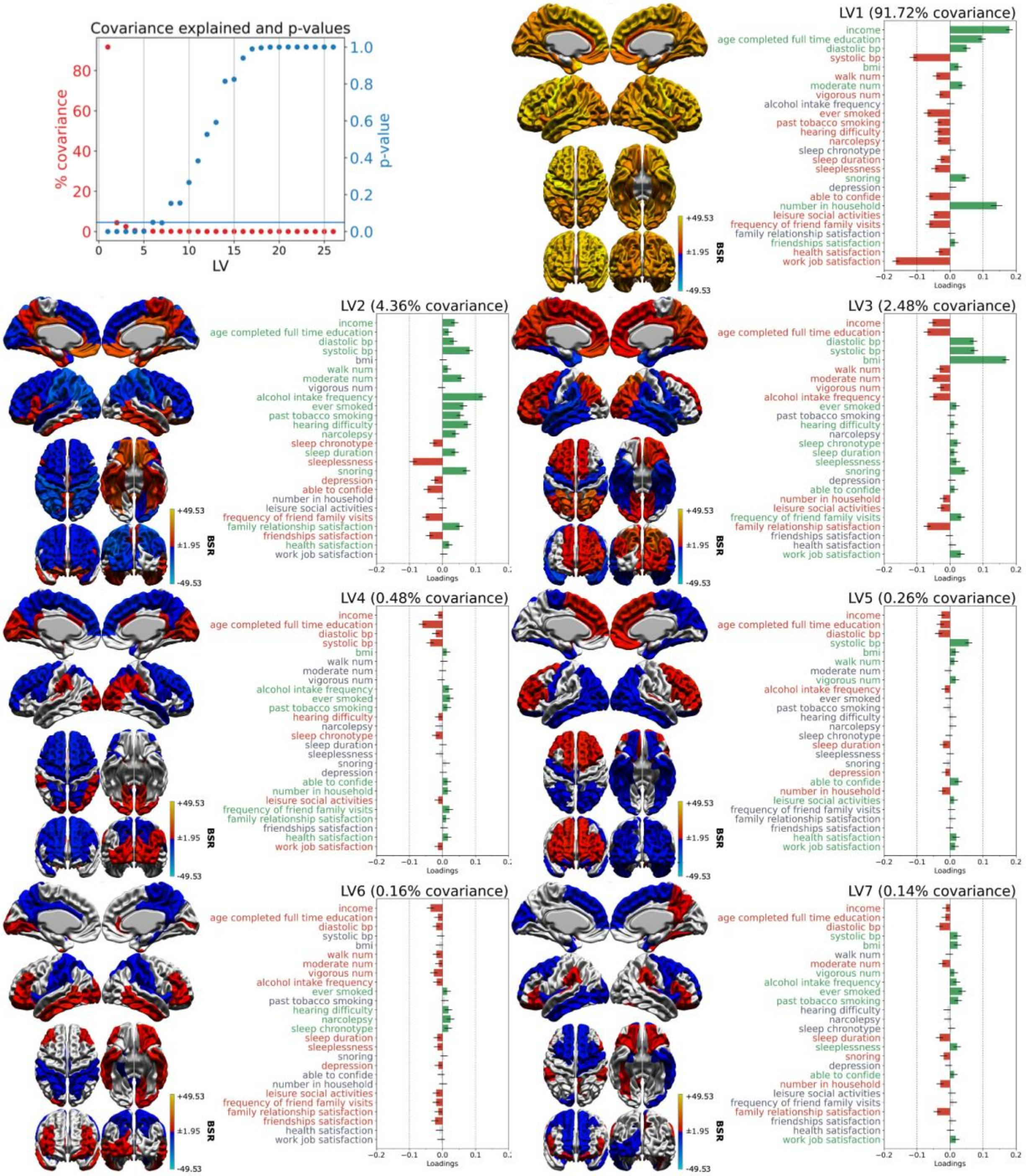


**Figure S1: Latent Variables of Whole Sample Analysis Figure 1:** PLS found 7 significant ($p < 0.05$) latent variables (LVs) in the whole sample analysis. Covariance explained (red) and significance (blue) of each latent variable in the top left. The horizontal blue line represents the significance threshold of p-values at 0.05. The remaining plots showcase the morphometric (left) and behavioral (right) patterns for each LV. Warm colors on morphometric maps indicate regions that vary positively with the LV, and cold collars indicate regions that vary negatively. Colored bars in behavioral loading plots indicate significant contributions to the clinical-behavioral signature, meaning maximal covariance with colored regions on the brain map. Green indicates a positive loading, and red indicates a negative loading. 95% confidence intervals are shown, and variables have non-significant contributions to the LV if the confidence interval crosses 0. Red indicates a positive loading, and blue indicates a negative loading. (**BSR**: bootstrap ratio)

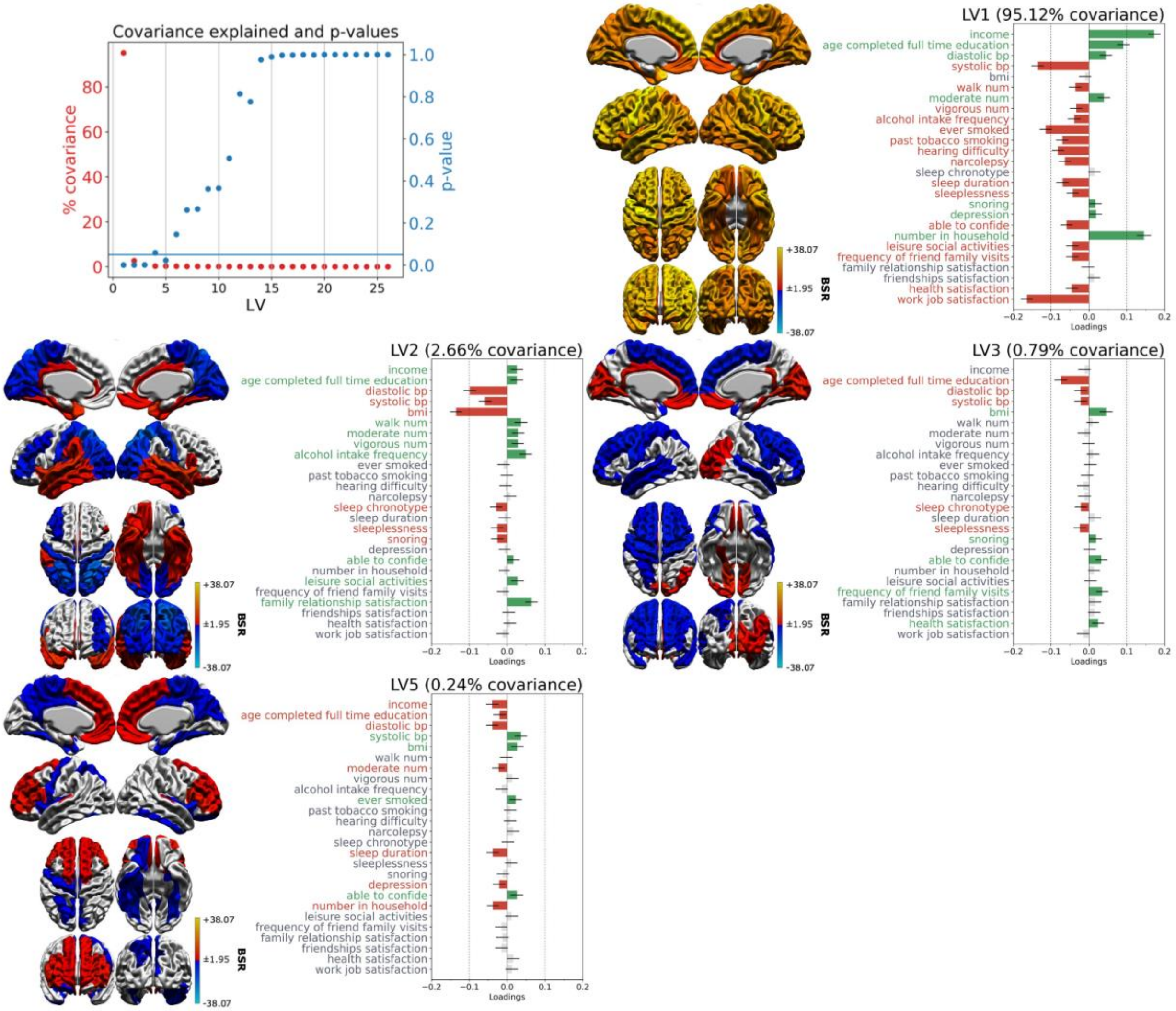


**Figure S2: Latent Variables of male-specific analysis:** PLS found 4 significant ($p < 0.05$) latent variables (LVs) in the male sample analysis. Covariance explained (red) and significance (blue) of each latent variable in the top left. The horizontal blue line represents the significance threshold of p-values at 0.05. The remaining plots showcase the morphometric (left) and behavioral (right) patterns for each LV. Warm colors on morphometric maps indicate regions that vary positively with the LV, and cold collars indicate regions that vary negatively. Colored bars in behavioral loading plots indicate significant contributions to the clinical-behavioral signature, meaning maximal covariance with colored regions on the brain map. Green indicates a positive loading, and red indicates a negative loading. 95% confidence intervals are shown, and variables have non-significant contributions to the LV if the confidence interval crosses 0. Red indicates a positive loading, and blue indicates a negative loading. (**BSR**: bootstrap ratio)

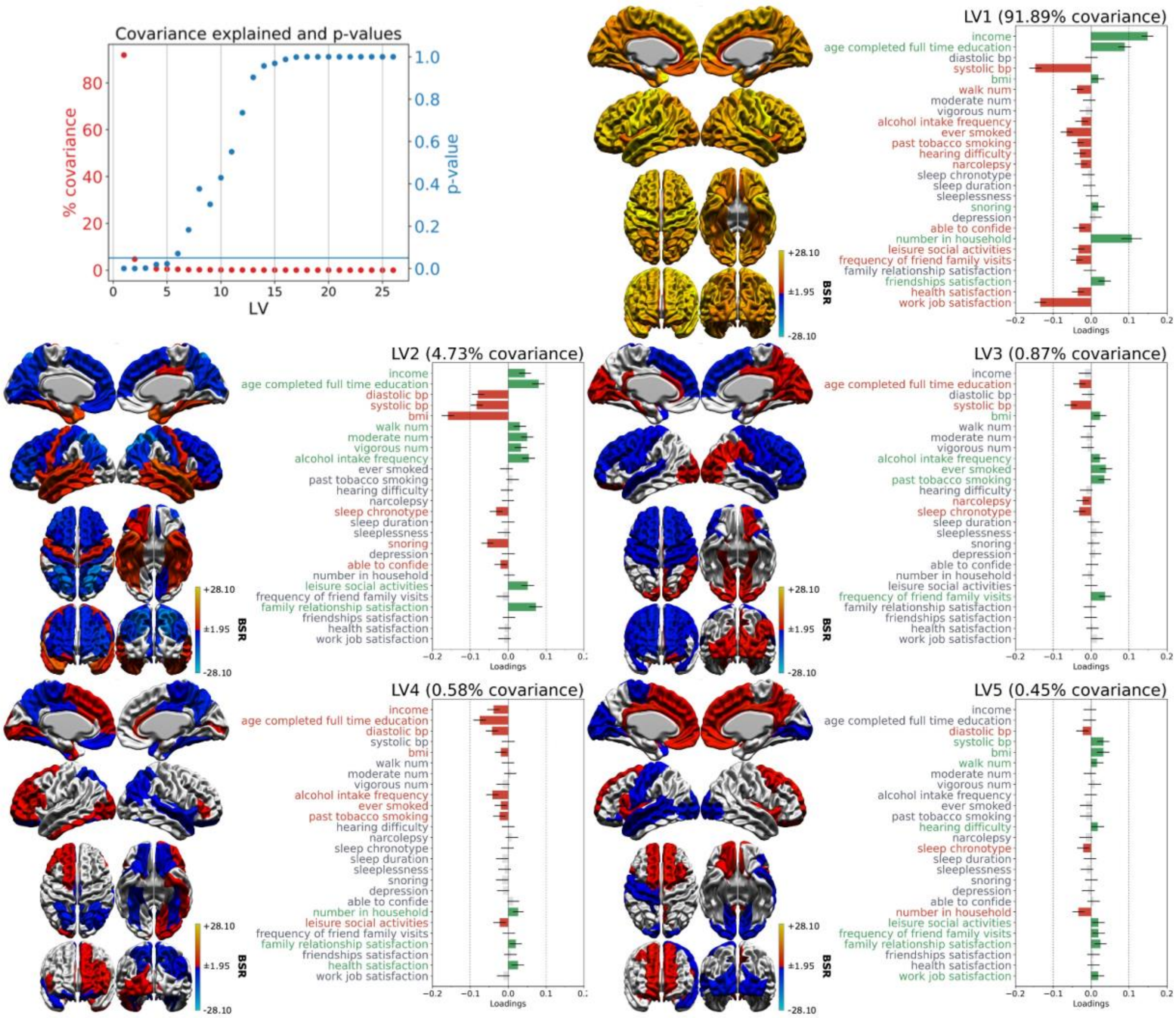


**Figure S3: Latent Variables of female-specific analysis** PLS found 5 significant ($p < 0.05$) latent variables (LVs) in the female analysis. Covariance explained (red) and significance (blue) of each latent variable in the top left. The horizontal blue line represents the significance threshold of p-values at 0.05. The remaining plots showcase the morphometric (left) and behavioral (right) patterns for each LV. Warm colors on morphometric maps indicate regions that vary positively with the LV, and cold collars indicate regions that vary negatively. Colored bars in behavioral loading plots indicate significant contributions to the clinical-behavioral signature, meaning maximal covariance with colored regions on the brain map. Green indicates a positive loading, and red indicates a negative loading.  95% confidence intervals are shown, and variables have non-significant contributions to the LV if the confidence interval crosses 0. Red indicates a positive loading, and blue indicates a negative loading. (**BSR**: bootstrap ratio)

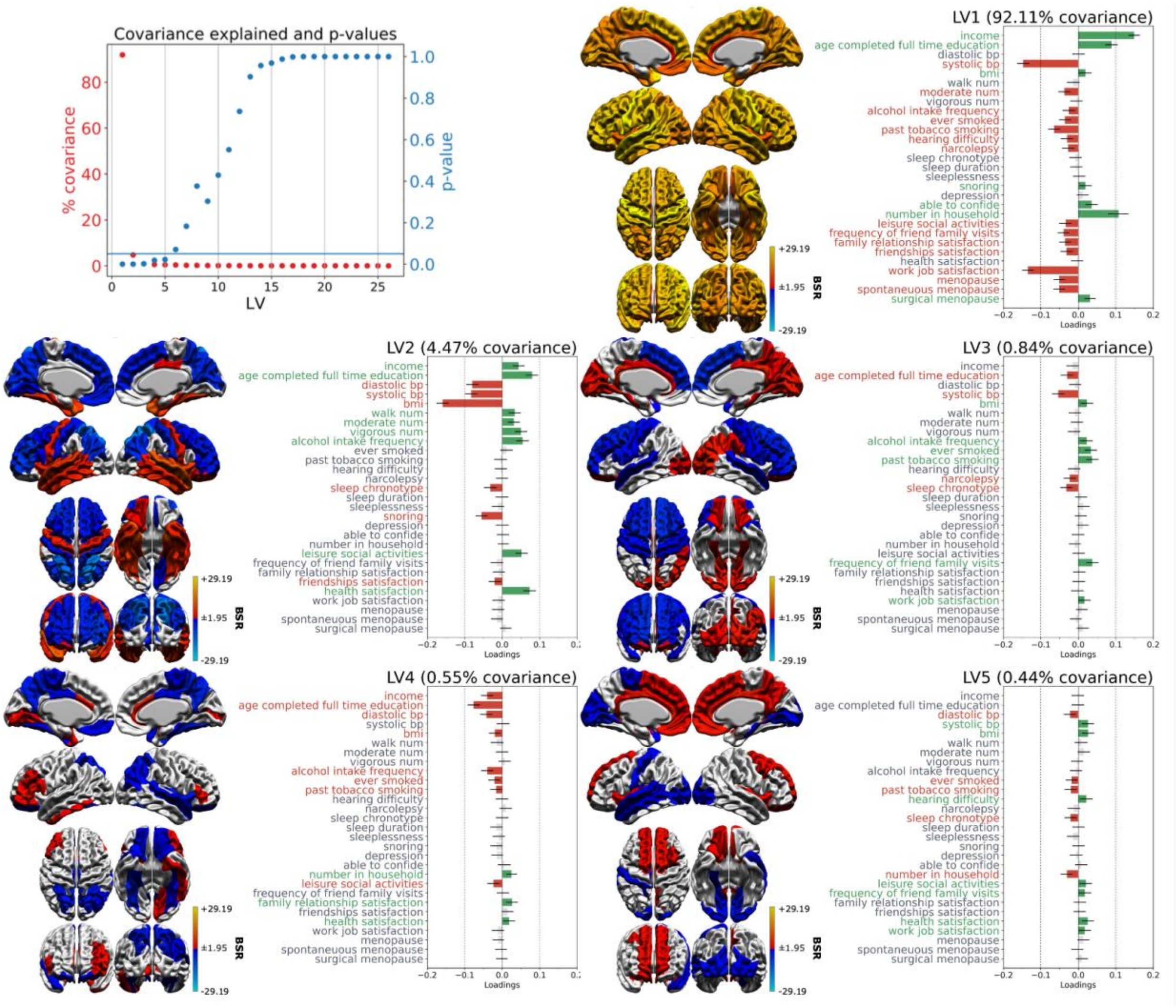


**Figure S4: Latent Variables of female-specific analysis with inclusion of menopause variables** PLS found 5 significant ($p < 0.05$) latent variables (LVs) in the female menopause-related analysis. Covariance explained (red) and significance (blue) of each latent variable in the top left. The horizontal blue line represents the significance threshold of p-values at 0.05. The remaining plots showcase the morphometric (left) and behavioral (right) patterns for each LV. Warm colors on morphometric maps indicate regions that vary positively with the LV, and cold collars indicate regions that vary negatively. Colored bars in behavioral loading plots indicate significant contributions to the clinical-behavioral signature, meaning maximal covariance with colored regions on the brain map. Green indicates a positive loading, and red indicates a negative loading. 95% confidence intervals are shown, and variables have non-significant contributions to the LV if the confidence interval crosses 0. Red indicates a positive loading, and blue indicates a negative loading. (**BSR**: bootstrap ratio)

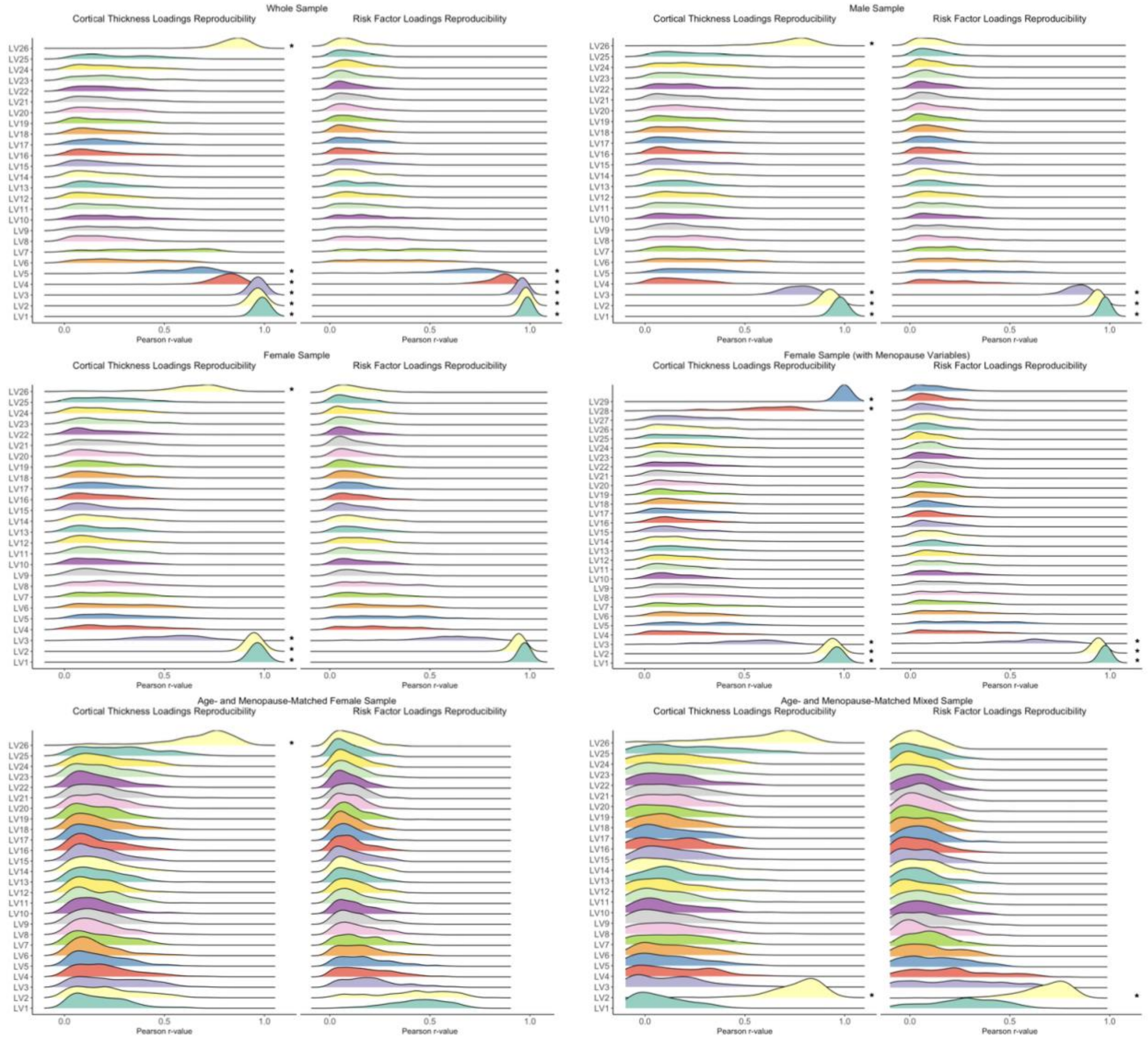


**Figure S5: Distributions from split-half stability analysis for each PLS model.** Split-half resampling analysis found varying levels of stability within each PLS model. In each plot, each row represents the distribution for Pearson correlation coefficients between respective loadings from each split half analysis. Distributions centered around zero indicate minimal correspondence between loadings from each split-half analysis, indicating higher sensitivity of participant characteristics.  Asterisks indicate the LVs variables (LVs) that showed a significant distribution (Z-score > 1.96).
